# Comparison of United Kingdom (UK) and United States (U.S.) hypertension treatment status, physical activity and prospective mortality risk

**DOI:** 10.64898/2026.03.25.26349345

**Authors:** Chenna Wang, Raaj K. Biswas, Nicholas A. Koemel, Matthew N. Ahmadi

## Abstract

**Background:** The 2017 American College of Cardiology/American Heart Association (ACC/AHA) guideline lowered diagnostic threshold for hypertension, encouraging earlier treatment initiation in the U.S. compared to UK, where the National Institute for Health and Care Excellence (NICE) guideline recommends higher thresholds. No comparative study evaluating how different hypertension guidelines and physical activity are jointly associated with mortality outcomes in two countries.

**Aims:** This study compared hypertension prevalence, treatment uptake, blood pressure (BP) levels, and mortality between the UK Biobank (UKBB) and the U.S. National Health and Nutrition Examination Survey (NHANES). We evaluated whether moderate-to-vigorous physical activity (MVPA) modifies mortality risk among different hypertension subgroups (normotensive, medicated hypertension, and unmedicated hypertension).

**Methods:** We harmonized demographic, biomarker, lifestyle, and accelerometer data from UKBB (n=63,452) and NHANES (n=7,397). Comprehensive weighting methods were applied in both cohorts. Accelerometry data was classified using a validated two-stage machine learning Random Forest algorithm. Associations between MVPA and all-cause mortality were examined with restricted cubic spline regression and visualized using Kaplan-Meier survival curves.

**Results:** NHANES showed a higher proportion of treated hypertension (29.9%) and lower average blood pressure (SBP/DBP: 122.2/70.7 mmHg) compared to UKBB (11.7% treated; SBP/DBP: 136.0/81.3 mmHg). Despite lower BP levels, cardiovascular mortality was higher in NHANES (10.3 per 10,000 person-years) compared to UKBB (4.0 per 10,000 person-years). In both cohorts, greater MVPA was linked to lower mortality risk, with the strongest association observed among medicated hypertensives. Notably, NHANES participants with treated hypertension and low MVPA (<10.7 minutes/day) experienced a steeper survival decline, falling to 74% by year 8, compared to 91% in normotensives and 79% in untreated hypertensives.

**Conclusion:** Despite higher treatment prevalence and lower average BP levels in NHANES, mortality remained higher compared with UKBB, suggesting that differences in mortality patterns may relate to broader cardiometabolic profiles and PA patterns beyond pharmacological management alone. Across both cohorts, higher levels of MVPA were associated with lower all-cause mortality, with the strongest associations were observed among individuals with medicated hypertension.

## Introduction

Hypertension is a leading modifiable risk factors for premature cardiovascular mortality and long-term disability worldwide, affecting an estimated 1.39 billion adults worldwide ^1–3^. Elevated BP is consistently associated with increased mortality risk from ischemic heart disease and stroke ^2, 4^. Effective BP control is therefore essential to alleviate the global cardiovascular disease (CVD) burden.

However, definitions of hypertension differ across national guidelines, influencing both prevalence estimates and treatment strategies. The 2017 ACC/AHA guideline lowered the diagnostic threshold for hypertension to BP ≥130/80 mm Hg, advocating earlier initiation of pharmacologic treatment ^5^. By contrast, the UK’s NICE guideline maintains a higher threshold of BP ≥140/90 mm Hg, resulting in more conservative management practices ^6^. These divergent thresholds raise critical questions regarding their respective impacts on hypertension detection, treatment uptake, and long-term cardiovascular outcomes. Evidence from the ESPRIT trial, shows that while intensive systolic blood pressure (SBP) lowering (<120 mm Hg) does not reduce total stroke incidence over 3.4 years, it can halve the risk of haemorrhagic stroke after about one year of treatment ^7^, underscoring how the benefits of tighter control depend on stroke subtype, patient characteristics, and the duration of exposure.

Physical activity (PA) has been widely recognized as a key nonpharmacological component in hypertension management. Regular PA lowers BP through improvements in vascular function and autonomic regulation, effects that tend to be more pronounced in people with hypertension ^8–11^. Combining regular PA with pharmacologic treatment produces synergistic benefits, substantially lowering cardiovascular risk ^9^. Despite these known relationships, a cross-national accelerometer study suggests that Americans, particularly those over age 50, are markedly less active than UK adults ^12^. These findings suggest important cross-national differences in PA patterns. Despite clear evidence supporting early intervention and regular PA for hypertension management, it remains unclear to what extent differences in national hypertension guidelines and PA patterns contribute to cardiovascular outcomes.

Compounding these differences, evidence indicates that cardiovascular mortality rates in the U.S. are substantially higher than those observed in other high-income countries, such as the UK ^13, 14^. Taken together, these findings indicate that lower treatment thresholds in the U.S. have not necessarily corresponded with better cardiovascular outcomes. This pattern may reflect broader differences in cardiometabolic risk profiles and PA patterns.

This study uses data from two large, population-based cohorts, UKBB and NHANES, to compare hypertension prevalence, treatment patterns, and mortality under unified hypertension criteria. The study aims to explore whether accelerometer-measured PA levels modify the association between hypertension status and mortality within each cohort.

## Methods

### Study Design and Data Sources

Our study utilizing existing data from two well-established population-based cohorts: UKBB and NHANES ^15–18^. These datasets include detailed demographic, behavioural, and clinical information, making them well-suited to evaluate how national hypertension guidelines and PA patterns influence mortality.

UKBB is a large, population-based cohort study established to investigate factors contributing to major diseases in middle-aged and older adults. It recruited 500,000 adults aged 40 to 69 years during 2006–2010 ^16, 19^. UKBB data are linked to longitudinal mortality records through NHS Digital for participants in England and Wales, and through the NHS Central Register for participants in Scotland ^20^. Wearables-based PA data were collected from a subset of 103,617 participants, each wearing triaxial accelerometers (Axivity AX3) on the wrist continuously for seven days at a sampling frequency of 100Hz ^16, 21^.

NHANES is a recurring national survey conducted by the U.S. Centres for Disease Control and Prevention (CDC) to assess the health and nutritional status of the non-institutionalized civilian population. In addition to cross-sectional data, NHANES is linked to longitudinal mortality follow-up through the National Death Index (NDI) ^22^. The linked mortality files provide detailed individual-level time-to-event data, including date and cause of death, duration of follow-up, and mortality status. During the 2011-2014 cycles, a subsample of participants wore triaxial accelerometers (ActiGraph GT3X+) on wrist for seven consecutive days, recording data at 80 Hz sampling frequency ^17, 23^.

### Eligibility Criteria and Data Extraction

We included participants aged 20 years or older if they have valid BP measurements, complete accelerometer data, and no missing values for key exposure and outcome variables. We conducted data harmonisation to align variable definitions and ensure consistency between UKBB and NHANES. We standardized variables using consistent definitions, measurement units, and clinically relevant cut-offs across cohorts. PA data derived from accelerometry was classified using a two-stage machine learning Random Forest algorithm, validated in prior studies ^24–30^. In the first stage, 10-second windows were assigned to one of four activity classes (sedentary, standing utilitarian movements, walking activities, or running/high energetic activities). They were then mapped to sedentary, light (<100 mg), moderate (≥100 mg), or vigorous (≥400 mg) intensity levels ^28–31^. The total duration of activity in each intensity band was aggregated across all valid wear days. Before conducting mortality analyses, we applied further exclusions to improve the validity of risk estimates. Participants with incomplete mortality data were removed. We excluded participants who rated their health as poor or fair to minimise confounding by baseline frailty ^28–30^. We excluded individuals who died within the first year of follow-up to reduce reverse causality risk ^28–30^.

### Statistical Analysis Plan

Participants were categorised into three mutually exclusive hypertension groups based on the 2017 ACC/AHA guideline: (1) normotensive (SBP <130 mm Hg and DBP <80 mm Hg without BP-lowering medications), (2) unmedicated hypertension (SBP ≥130 mm Hg or DBP ≥80 mm Hg without BP-lowering medications), and (3) medicated hypertension (self-reported use of antihypertensive medications) ^5^. This classification was applied uniformly across both datasets, facilitating direct comparisons between cohorts. We selected the 2017 ACC/AHA guideline because it marked a paradigm shift in hypertension management in the U.S., while most other countries maintain the long-standing benchmark of 140/90 mmHg ^5, 32^. Despite different diagnostic thresholds, there is strong international agreement that individuals at high atherosclerotic cardiovascular disease (ASCVD) risk should aim for a BP target of <130/80 mmHg ^33^.

To mitigate the influence of extreme outliers, we winsorized moderate-to-vigorous physical activity (MVPA) and measurements of SBP and DBP at the 2.5^th^ and 97.5^th^ percentiles ^24, 26, 28–30^. We applied robust weighting methods on both cohorts ^34–36^. For UKBB, we applied inverse probability weighting (IPW) based on recent work by van Alten et al. to correct for selection bias due to non-random volunteer participation ^37^. For NHANES, we incorporated sampling weights provided by the National Centre for Health statistics (NCHS) to produce unbiased, population-level estimates representative of the non-institutionalized U.S. civilian population ^17^.

We used Cox proportional hazards regression models to examine the relationship between MVPA tertiles and all-cause mortality, separately within each hypertension group in both cohorts ^28–30, 38^. Covariates included age, sex, ethnicity, smoking status, alcohol intake, fruit and vegetable consumption, educational attainment, previous CVD, family history of CVD, sleep duration, and light physical activity (LPA) energy expenditure. We used forest plots to present the hazard ratios (HRs) and 95% confidence intervals (CIs) for the intermediate and high MVPA tertiles relative to the low MVPA reference group. Additionally, we generated covariate-adjusted Kaplan-Meier survival curves to estimate survival probabilities across hypertension subgroups stratified by MVPA tertiles within each cohort.

All statistical analyses were conducted using R (version 4.4.1).

### Subgroup and sensitivity analysis

To further validate the robustness of the primary findings, we conducted several subgroup and sensitivity analyses. First, given the differing age distributions between cohorts, we restricted the NHANES sample to adults aged 40–69 years to match the UKBB age range (Supplementary Figure 5–6). Second, we excluded participants with self-reported cardiovascular disease at baseline to minimise reverse causation (Supplementary Figure 7–8) ^28–30, 38^. To address the markedly higher baseline MVPA levels observed in UKBB compared with NHANES, we applied NHANES-derived MVPA tertile thresholds uniformly to both datasets (Supplementary Figure 9). This allowed direct comparison of survival outcomes at comparable MVPA levels. In addition, we conducted a sensitivity analysis using unweighted data from UKBB and NHANES to o evaluate whether the application of weighting methods in the primary analyses materially altered the observed survival patterns (Supplementary Figure 10).

## Results

### Participants demographics and characteristics

The weighted baseline demographic, behavioural, and clinical characteristics of participants from UKBB and NHANES are presented in Supplementary Table 1, while the unweighted characteristics are shown in Supplementary Table 2. The UKBB sample included 63,452 participants with a mean age of 58.2 years (SD = 8.3) and the NHANES sample consisted of 7,397 individuals, who were younger (mean age = 49.3 years, SD = 16.6). The ethnic composition also varied markedly: over 94% of UKBB participants identified as White, whereas NHANES included approximately 68% White, 11% Black, 4% Asian, and 16% Other (Supplementary Table 1; Supplementary Figure 1).

Lifestyle differences between cohorts were also evident. For example, PA patterns further highlighted disparities: non-exercisers were more common in NHANES (46.4%) than in UKBB (32.2%), and daily duration of MVPA was approximately double in UKBB (40.0 ± 25.2 min/day) compared to NHANES (21.0 ± 17.4 min/day). Average daily alcohol intake was moderately higher in NHANES (2.6 ± 2.2 units/day) compared to UKBB (1.9 ± 2.3 units/day). NHANES participants had higher average waist circumference and body mass index (BMI) (100.2 cm and 29.3 kg/m², respectively) relative to their UKBB counterparts (88.3 cm and 26.7 kg/m²). Despite these indicators of higher cardiometabolic risk in NHANES, SBP and DBP readings were paradoxically lower than in UKBB. However, this apparent advantage in BP did not translate into survival benefit. Cardiovascular mortality was more than three times higher in NHANES (2.3%) than in UKBB (0.7%; Supplementary Table 1, Figure 1).

**Figure 1:**
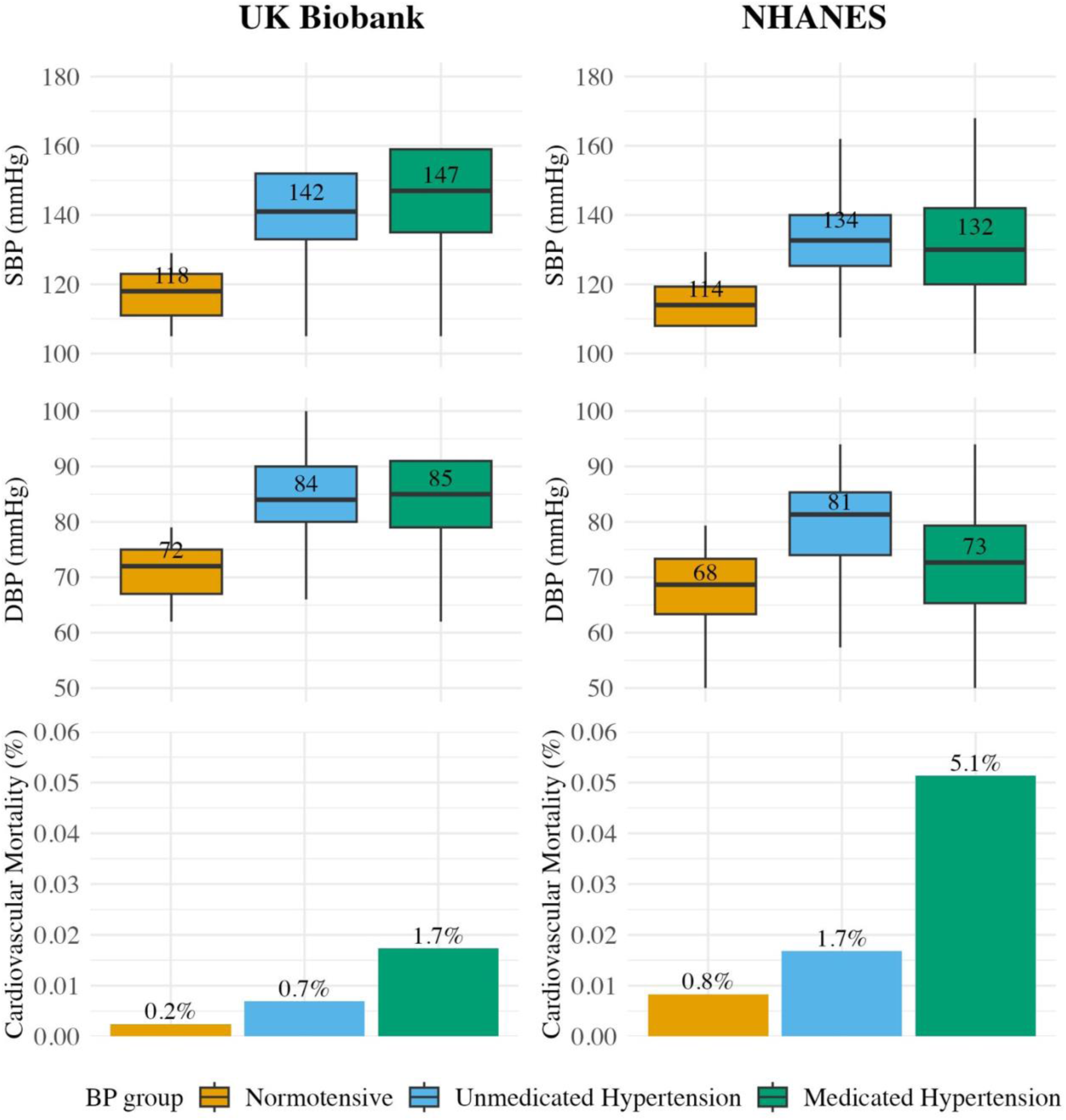
Weighted distribution of systolic blood pressure, diastolic blood pressure, and cardiovascular mortality by hypertension groups in UK Biobank and NHANES *Box plots show median SBP and DBP values across normotensive, unmedicated hypertensive, and medicated hypertensive subgroups in UKBB and NAHNES, including adults aged 20 years and older. Bar plots represent cardiovascular mortality rates within each hypertension subgroup*.

Across both cohorts, participants with medicated hypertension showed more adverse cardiometabolic profiles than their unmedicated hypertensive counterparts. These individuals were older (UKBB: 63.9 vs. 58.6 years; NHANES: 60.8 vs. 50.5 years) and more metabolically compromised in terms of HbA1C, HDL cholesterol, and triglycerides profiles. PA levels were also lower in this group. In UKBB, mean MVPA was 33.4 minutes per day compared to 40.1 minutes among unmedicated hypertensives. In NHANES, the gap was larger (13.8 vs. 22.9 minutes per day). Cardiovascular mortality was highest among medicated hypertensives in both datasets (UKBB: 1.7%, NHANES: 5.1%, Figure 1). These trends suggest that lower PA levels may compound the residual risk despite pharmacological treatment in medicated hypertension group.

### BP Profiles and Treatment Prevalence

The distribution of hypertension status under the 2017 ACC/AHA guideline differed substantially between cohorts. In UKBB, 29.7% of participants were normotensive, 58.6% had unmedicated hypertension, and 11.7% were receiving antihypertensive treatment. NHANES presented a different pattern: 50.7% were normotensive, 19.4% had unmedicated hypertension, and 29.9% were treated (Supplementary Figure 2). These differences suggest that U.S. adults were more likely to be diagnosed and treated with antihypertensives, prompted by the lower treatment threshold.

### All-cause mortality analysis

#### Cox analysis

In multivariable-adjusted models, higher levels of MVPA were associated with lower all-cause mortality risk in participants with medicated hypertension in UKBB when compared with the low MVPA tertile. In NHANES, only the intermediate MVPA tertile was significantly associated with lower mortality risk in medicated hypertension group (Supplementary Figure 4). Compared with the low MVPA tertile, the high MVPA tertile was associated with a HR of 0.60 (95% CI 0.41–0.88) in UKBB and 0.48 (95% CI 0.21–1.11) in NHANES, while the intermediate MVPA tertile was associated with an HR of 0.69 (95% CI 0.49–0.98) in UKBB and 0.47 (95% CI 0.24–0.94) in NHANES. In the unmedicated hypertension and normotensive subgroup, no statistically significant associations were observed. Across most hypertension categories, point estimates generally indicated lower mortality risk with higher MVPA, except in NHANES normotensives. Within NHANES, medicated hypertension had relatively narrower CIs than the other two hypertension groups because its total number of events (n = 260) was more than three times higher than in unmedicated hypertension (n = 78) or normotensives (n = 78). In the NHANES medicated hypertension group, the high MVPA tertile had a hazard ratio of 0.48 with a wider 95% CI of 0.21 to 1.11, likely due to having fewer deaths events (n = 26), less than half the number in the intermediate tertile (n = 53).

#### Kaplan–Meier survival curves

Covariate-adjusted Kaplan–Meier survival curves were used to depict the cumulative incidence of all-cause mortality over time across three hypertension groups, stratified by MVPA tertiles, in UKBB and NHANES (Figure 2). In UKBB, survival probabilities remained above 90% throughout the follow-up period across all MVPA strata, with minimal divergence between hypertension groups. However, medicated hypertensive participants in the low MVPA tertile showed an earlier and slightly steeper decline in survival probability compared to their normotensive and unmedicated hypertensive counterparts, while higher MVPA tertiles showed convergence of survival curves across hypertension groups.

**Figure 2:**
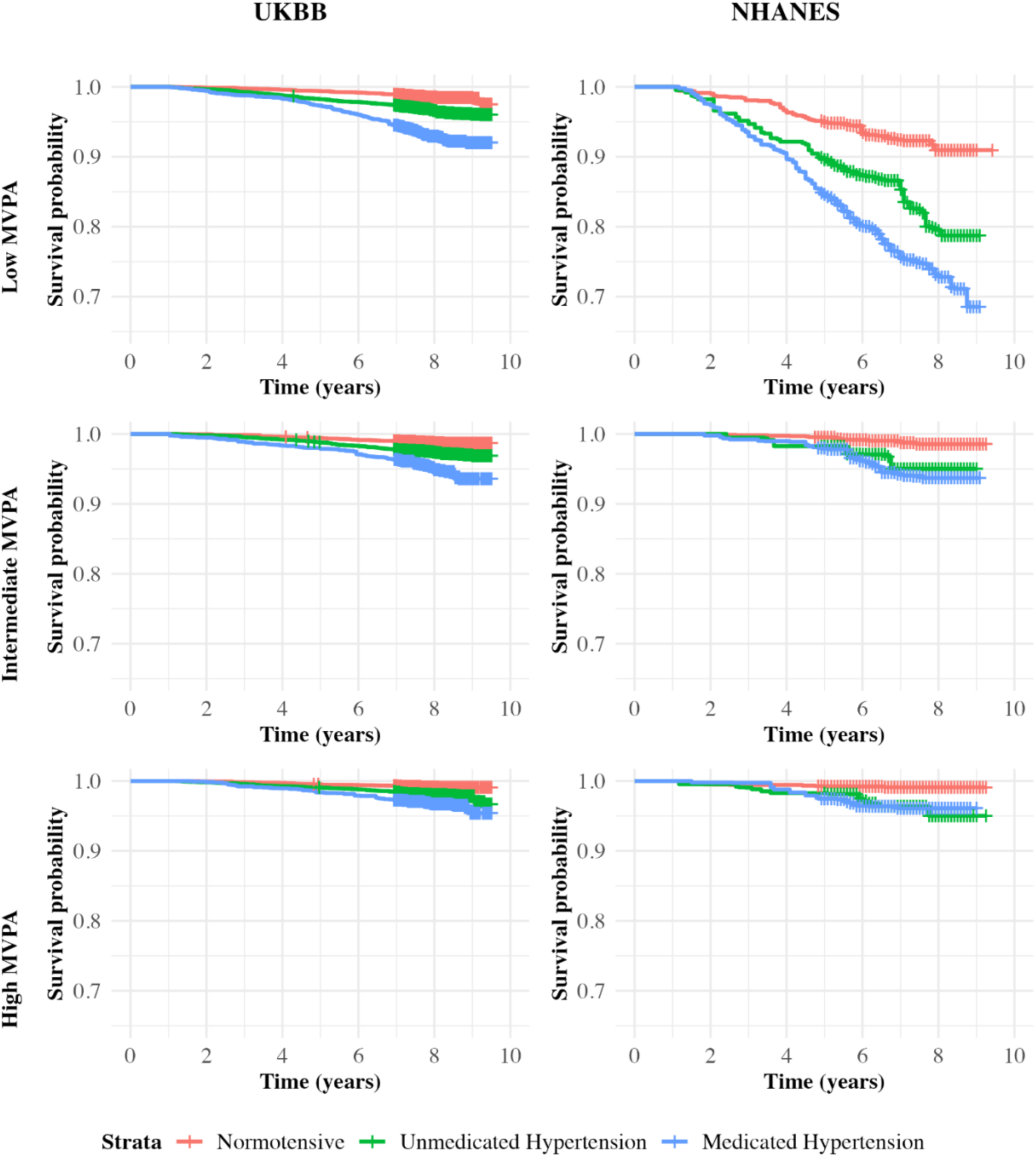
Weighted Kaplan-Meier survival curves for all-cause mortality by hypertension groups stratified by moderate-to-vigorous physical activity tertiles in UKBB and NHANES *MVPA tertiles were defined using cohort-specific distributions of accelerometer-measured daily duration of MVPA (minutes/day). For UKBB: Low (5.50–24.71), Intermediate (24.72–45.89), and High (≥45.90). For NHANES: Low (0.29–10.72), Intermediate (10.73–24.78), and High (≥24.79). Analysis included participants aged 20 years and older in UKBB and NHANES*.

In contrast, NHANES displayed clear separation of survival curves among hypertension groups, most evident in participants in low MVPA tertile. Medicated hypertensives in this group showed a steep decline in survival probability beginning early in the follow up period, reaching approximately 74% by year 8. This was substantially lower than normotensive and unmedicated hypertensive groups, which retained higher probabilities closer to 91% and 79%, respectively. However, these disparities were less evident in the intermediate and high MVPA strata, where survival curves for all groups converged above 90% at year 8. These patterns suggest that higher MVPA are associated with a lower mortality risk among individuals receiving antihypertensive treatment in both cohorts.

#### Sensitivity analysis

Sensitivity analyses supported the robustness of the main findings. Aligning NHANES to the UKBB age range and excluding participants with baseline cardiovascular disease did not materially alter the associations between MVPA, hypertension status, and survival. Applying NHANES-derived MVPA tertiles to both datasets provided clearer separation of survival curves at lower activity levels, supporting the interpretation that increased MVPA mitigates mortality risks across hypertension strata. In addition, analyses using unweighted data yielded Kaplan–Meier survival curves that were largely consistent with those from the weighted primary analyses.

## Discussion

### Key findings

The observed differences in hypertension status between UKBB and NHANES are closely linked to distinct diagnostic and treatment criteria set by NICE and ACC/AHA guidelines. Specifically, the ACC/AHA 2017 guideline defines hypertension at a lower threshold. It defines stage 1 hypertension as SBP 130–139 mmHg or DBP 80–89 mmHg and recommends medication for adults within this range if their estimated 10-year ASCVD risk is 10% or higher ^5^. By contrast, treatment under NICE typically starts at clinic BP ≥160/100 mmHg or ≥140/90 mmHg when significant cardiovascular risk factors or target organ damage exist ^6^.

Despite lower mean BP among medicated hypertensive participants in NHANES (130.8/71.0 mmHg) compared with UKBB (147.2/85.8 mmHg), cardiovascular mortality remained higher in NHANES (5.1% vs 1.7%). One potential explanation for the difference in cardiovascular mortality rates involves differences in overall cardiometabolic health profiles. NHANES participants exhibited poorer glycaemic control (5.7% vs. 5.4% in UKBB), lower HDL (52.9 mg/dL vs. 56.4 mg/dL), and greater obesity burden as indicated by waist circumference (100.2 cm vs. 88.3 cm) and BMI (29.3 kg/m² vs. 26.7 kg/m²). Additionally, NHANES participants showed less favourable lifestyle behaviours overall, particularly less daily MVPA (21.0 vs. 40.0 minutes/day). MVPA is a key lifestyle factor due to its robust and well-established relationship with cardiovascular and all-cause mortality ^39–41^. MVPA can be objectively measured, allowing for precise evaluation of its health impact. Therefore, this study focused on examining MVPA’s role in mitigating mortality risk in hypertension subgroups.

Higher MVPA levels were associated with lower all-cause mortality risk, especially among medicated hypertensive participants in both cohorts. Compared with the low MVPA tertile, the intermediate tertile was associated with a HR of 0.69 (95% CI: 0.49–0.98) in UKBB and 0.47 (95% CI: 0.24–0.94) in NHANES. Covariate-adjusted Kaplan-Meier survival curves supported this, with NHANES medicated hypertensive individuals in the lowest MVPA tertile (0.3–10.7 minutes/day) experiencing significantly lower survival probabilities (∼74% at year 8), compared to over 90% survival in the intermediate (10.8–24.8 minutes/day) and high (≥24.8 minutes/day) MVPA tertiles. These findings indicate that the association between MVPA and mortality varies by treatment status, with the effect most evident among medicated hypertensive individuals. Regular PA modulates the renin-angiotensin-aldosterone system, thereby improving BP control ^10, 11^. PA also reduces sympathetic tone, lowers peripheral vascular resistance, and enhances vascular function ^42^. Together, these mechanisms may help contextualize the observed associations between MVPA and lower mortality among medicated hypertensive participants.

We observed no statistically significant association between MVPA and all-cause mortality among normotensive participants. This observation aligns with prior findings that individuals who are already healthy may experience smaller measurable benefits from increased PA compared with those at higher risk ^43^. To better understand this result, we examined the distribution of mortality causes in this subgroup. Among all recorded deaths, only 15.9% were attributable to cardiovascular causes, while 59.5% were attributed to cancer. This suggests that in normotensive individuals, cancer-related mortality may predominate and dilute the overall association between MVPA and all-cause mortality.

### Clinical implication

Our findings emphasize the insufficiency of antihypertensive medication alone in mitigating mortality risk, particularly when other cardiometabolic risk factors or unhealthy lifestyle behaviours persist. Recognizing this complexity, clinicians should adopt a comprehensive, multifactorial approach that concurrently targets BP regulation, glycaemic and lipid optimisation, and sustained lifestyle modifications.

Among modifiable risk factors, intensive lifestyle interventions can significantly lower cardiovascular risk ^44, 45^. Among the lifestyle factors, our study highlights the pronounced mortality benefit of increased MVPA among medicated hypertensive patients. Regular PA not only lowers BP ^46, 47^, but also is associated with lower risk of excessive weight gain, cardiovascular disease, and all-cause mortality across diverse populations, including adults with preexisting chronic condition ^48, 49^. Even modest increases in activity can produce significant health benefits among the least active individuals ^48^. Our findings reinforce this by showing that among medicated hypertensives in NHANES, increasing MVPA from low to intermediate tertile was associated with an increase in 8-year survival from approximately 74% to over 90%. Clinicians should routinely prescribe PA alongside medications, supported by structured counselling and multidisciplinary teams ^50^. U.S. adults exhibit a higher burden of cardiometabolic risk factors alongside wider socioeconomic inequalities than their British counterparts ^51, 52^. These disparities are partly driven by systemic differences in healthcare structure and funding ^53–55^. These system-level distinctions highlight that optimal hypertension management requires not only clinical and behavioural interventions but also policies that address affordability of primary care and social infrastructure. Among these public health policies, this study draws particular attention to the social determinants that shape PA. Individual efforts must be supported by community infrastructure and accessible facilities. Neighbourhoods with connected street networks, accessible public transport, and plentiful parks led to significantly higher PA levels across diverse cities ^56^. Therefore, integrated, system-level interventions connecting clinical recommendations to supportive community environments can more effectively sustain behavioural changes and ultimately improve population health.

### Strength and limitation

Our study’s strengths include the use of wearable devices to objectively measure PA. Extensive evidence shows that self-reported measures provide only crude estimates of activity levels due to recall and reporting biases, whereas wearables provide more objective estimates on intensity and duration ^57, 58^. Furthermore, we used aligned analytic strategies across two large, independent population-based cohorts, facilitating direct comparison between UKBB and NHANES. The availability of rich demographic, lifestyle, and clinical data in both cohorts enabled comprehensive covariate adjustment, supporting more robust mortality analysis.

Our study also had several limitations. The one-week accelerometer measurement period might not adequately capture long-term patterns, especially given that PA behaviours fluctuate across the life course ^59^. Second, in UKBB, biomarker and accelerometry data were obtained approximately five years apart (baseline assessment in 2006–2010 and accelerometer measurements in 2013–2015). This temporal gap introduces potential inconsistencies, as participants’ hypertension profiles, medication use, or PA patterns could have changed during this period. Third, we lacked information on participants’ adherence to prescribed medications, which may have influenced the observed associations, especially when comparing medicated and unmedicated hypertension groups. Fourth, BP readings were taken once and cannot confirm chronic hypertension status, distinguish between controlled and uncontrolled hypertension within the medicated group, or assess individual trajectories of BP over time. In addition, we noted wider CIs in NHANES compared with UKBB, particularly in the unmedicated hypertension and normotensive subgroups. This pattern likely reflects substantially fewer all-cause mortality events in NHANES compared with UKBB (Supplementary Table 3), which diminished statistical power and amplified uncertainty in HR estimates. Future studies should aim to obtain long-term accelerometry data, access general practice records to assess medication adherence and repeated BP measurements, and include data from broader settings, particularly from low- and middle-income countries and datasets with greater volumes of mortality events.

## Conclusion

In conclusion, our cross-cohort comparative analysis highlights that higher levels of MVPA were consistently associated with lower all-cause mortality risks, with the most pronounced associations observed among individuals already receiving antihypertensive treatment. Despite higher rates of antihypertensive medication use and lower average BP readings in NHANES, cardiovascular mortality remained higher compared to UKBB. These findings suggest that variation in mortality patterns may reflect broader cardiometabolic profiles and PA patterns beyond pharmacological hypertension treatment alone. The results provide context for public health initiatives and clinical guidelines to consider how PA counselling and pharmacological management may jointly relate to long-term mortality outcomes across different population settings.

## List of abbreviations

ACC/AHA: (American College of Cardiology/American Heart Association)
ASCVD: (atherosclerotic cardiovascular disease)
BMI: (body mass index)
BP: (blood pressure)
CDC: (Centers for Disease Control and Prevention)
CI: (confidence interval)
CMD: (cardiometabolic diseases)
CVD: (cardiovascular disease)
DBP: (diastolic blood pressure)
HbA1c: (hemoglobin A1c)
HDL: (high-density lipoprotein cholesterol)
HR: (hazard ratio)
IPW: (inverse probability weighting)
LPA: (light physical activity)
MVPA: (moderate-to-vigorous physical activity)
NCHS: (National Center for Health Statistics)
NDI: (National Death Index)
NHANES: (National Health and Nutrition Examination Survey)
NHS: (National Health Service)
NICE: (National Institute for Health and Care Excellence)
PA: (physical activity)
RF: (random forest)
SBP: (systolic blood pressure)
SD: (standard deviation)
UK: (United Kingdom)
UKBB: (UK Biobank)
U.S.: (United States).

## Declaration

### Ethic approval and consent to participate

In UKBB, ethical approval for data collection was obtained from the North West Multi-Centre Research Ethics Committee. The NHANES protocol was approved by the US NHANES institutional review board and National Center for Health Statistics Research ethics review board. All participants provided written informed consent.

### Consent for publication

Not applicable.

### Availability of data and materials

NHANES data are available at www.cdc.gov/nchs/nhis/index.htm, and data from UK Biobank are available on application at www.ukbiobank.ac.uk/register-apply.

### Author’s contributions

CW, RKB, NAK, and MNA were major contributors to the study design, data analysis, drafting of the manuscript, and manuscript revision. All authors read and approved the final manuscript.

## Supplementary material

**Supplementary Figure 1:**
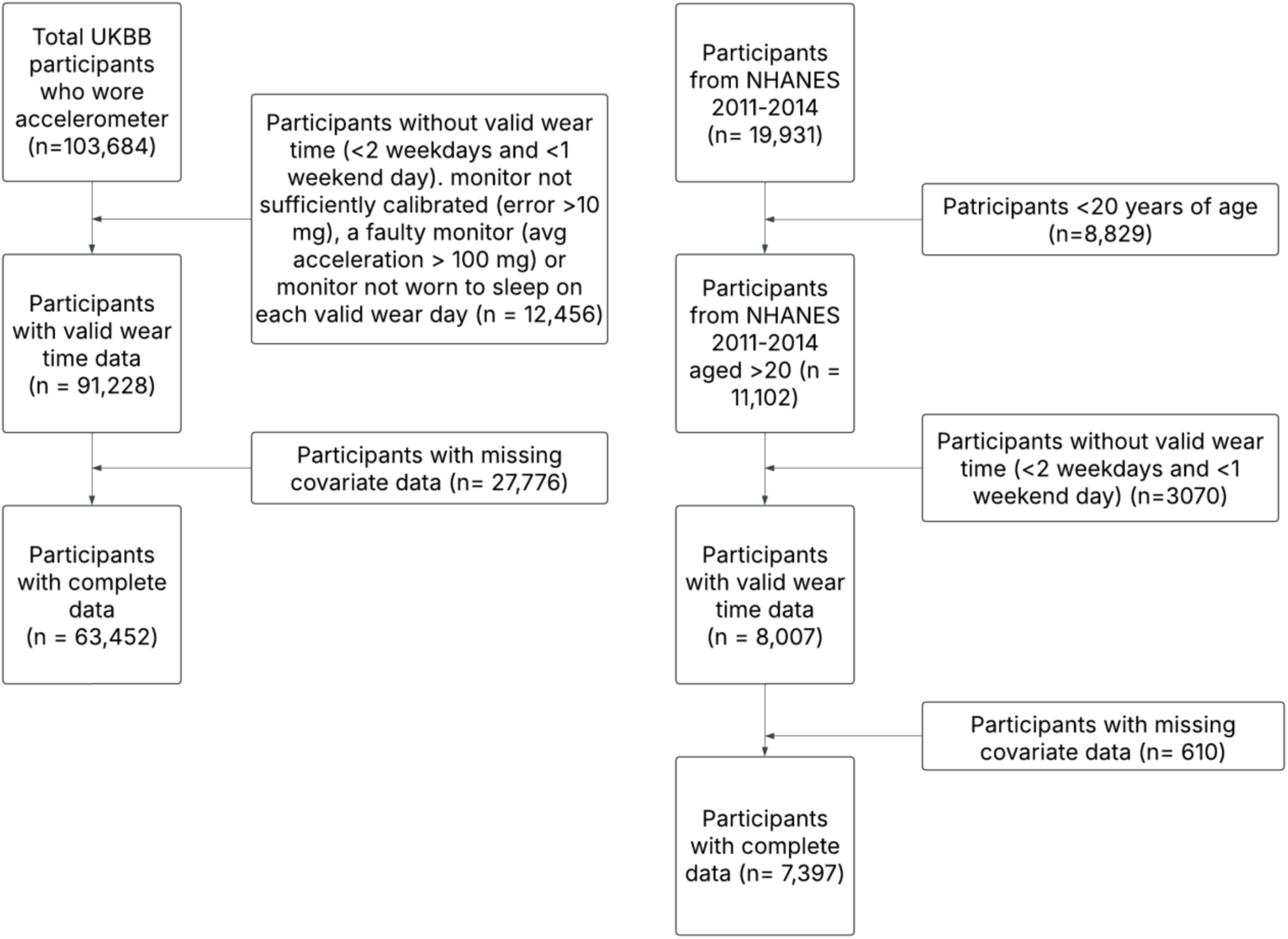
Flow diagram of participants in the study.

**Supplementary Figure 2:**
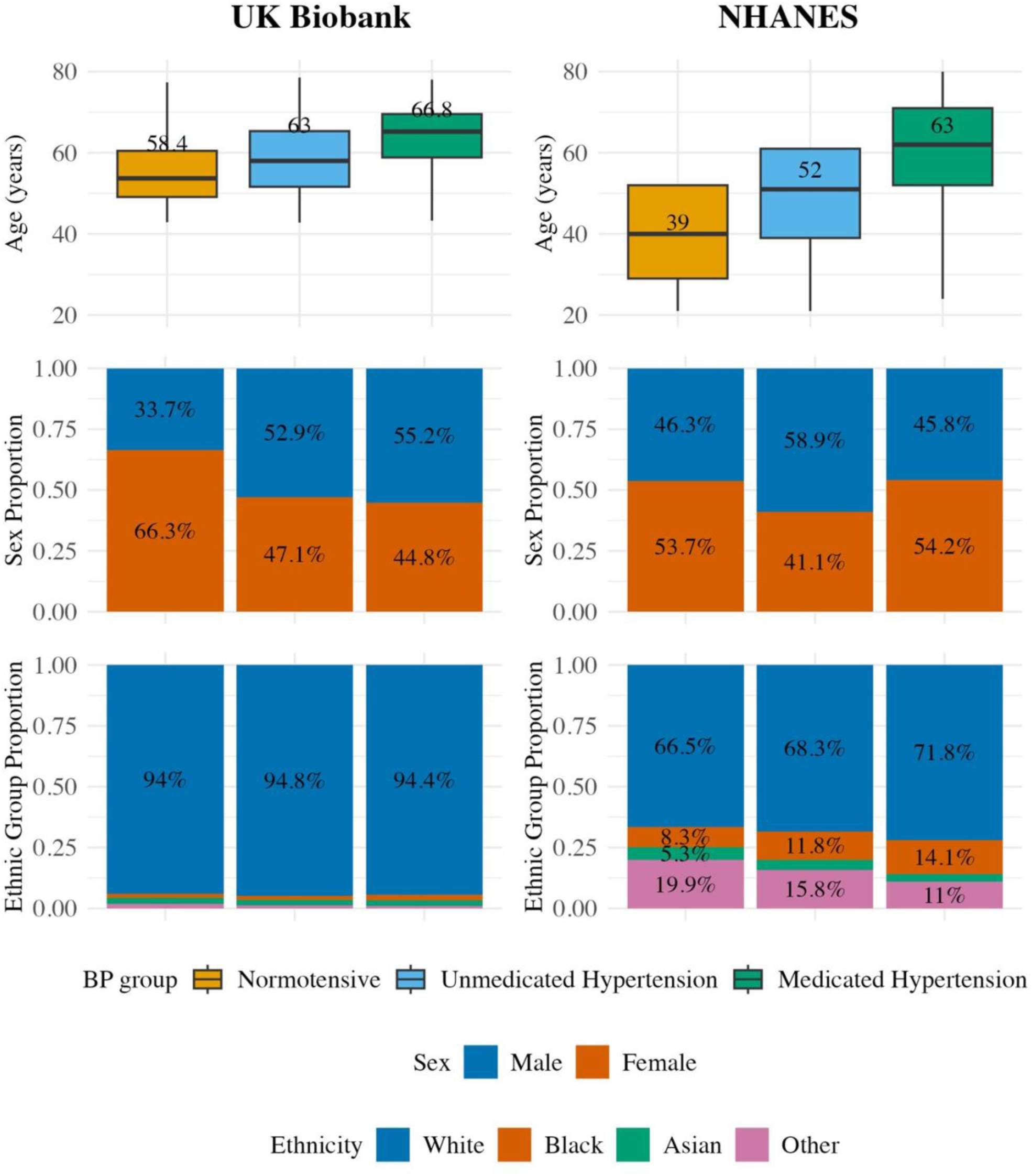
Weighted distribution of demographic factors by hypertension groups in UK Biobank and NHANES *Box plots display median age and interquartile ranges, while bar plots show sex and ethnic group proportions across hypertension subgroups within UKBB and NHANES*.

**Supplementary Figure 3:**
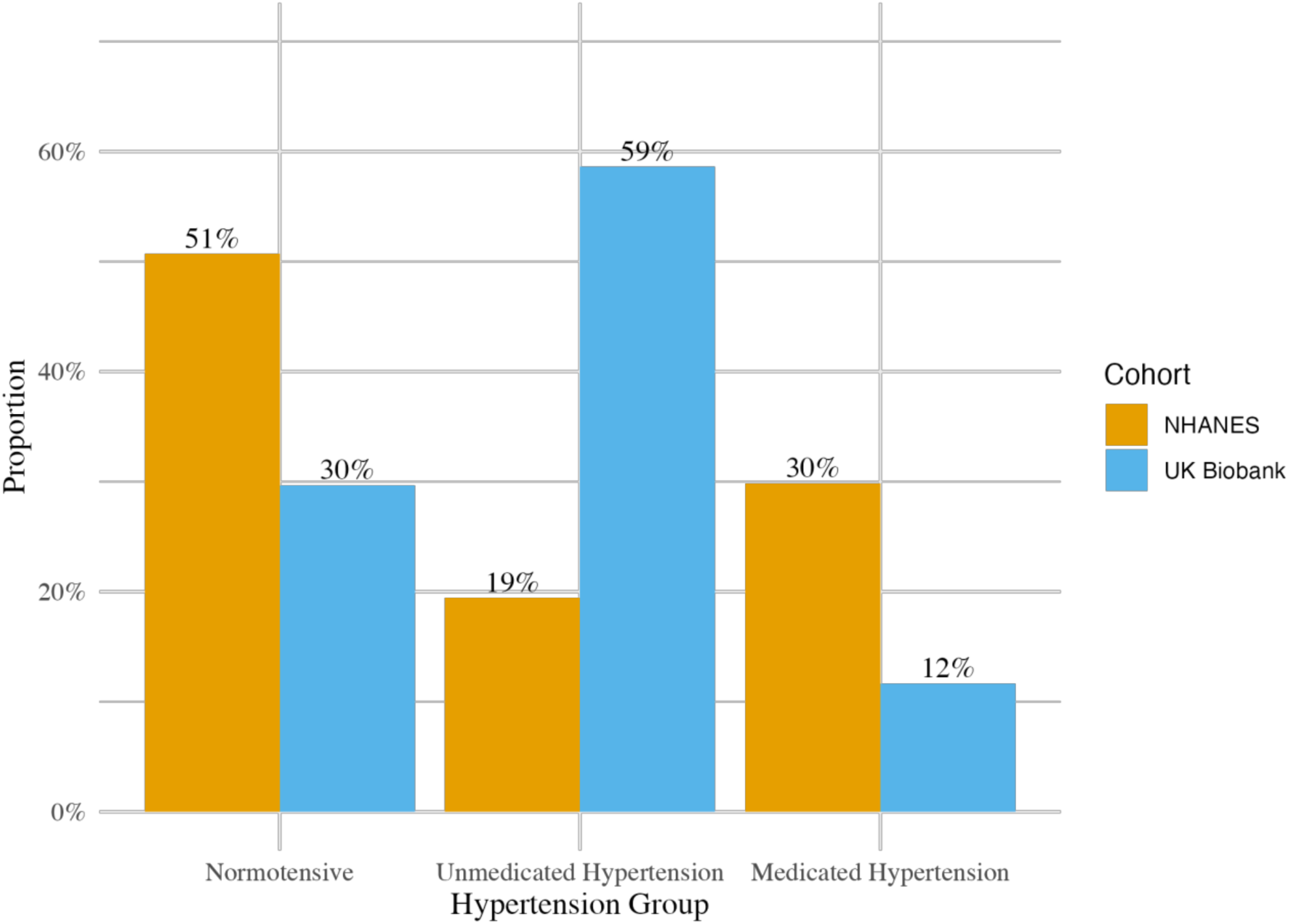
Weighted distribution of hypertension groups in UK Biobank and NHANES under the 2017 American College of Cardiology/American Heart Association guideline *Bar plots depict the percentage of normotensive, unmedicated hypertensive, and medicated hypertensive participants in each cohort. Values are weighted*.

**Supplementary Figure 4:**
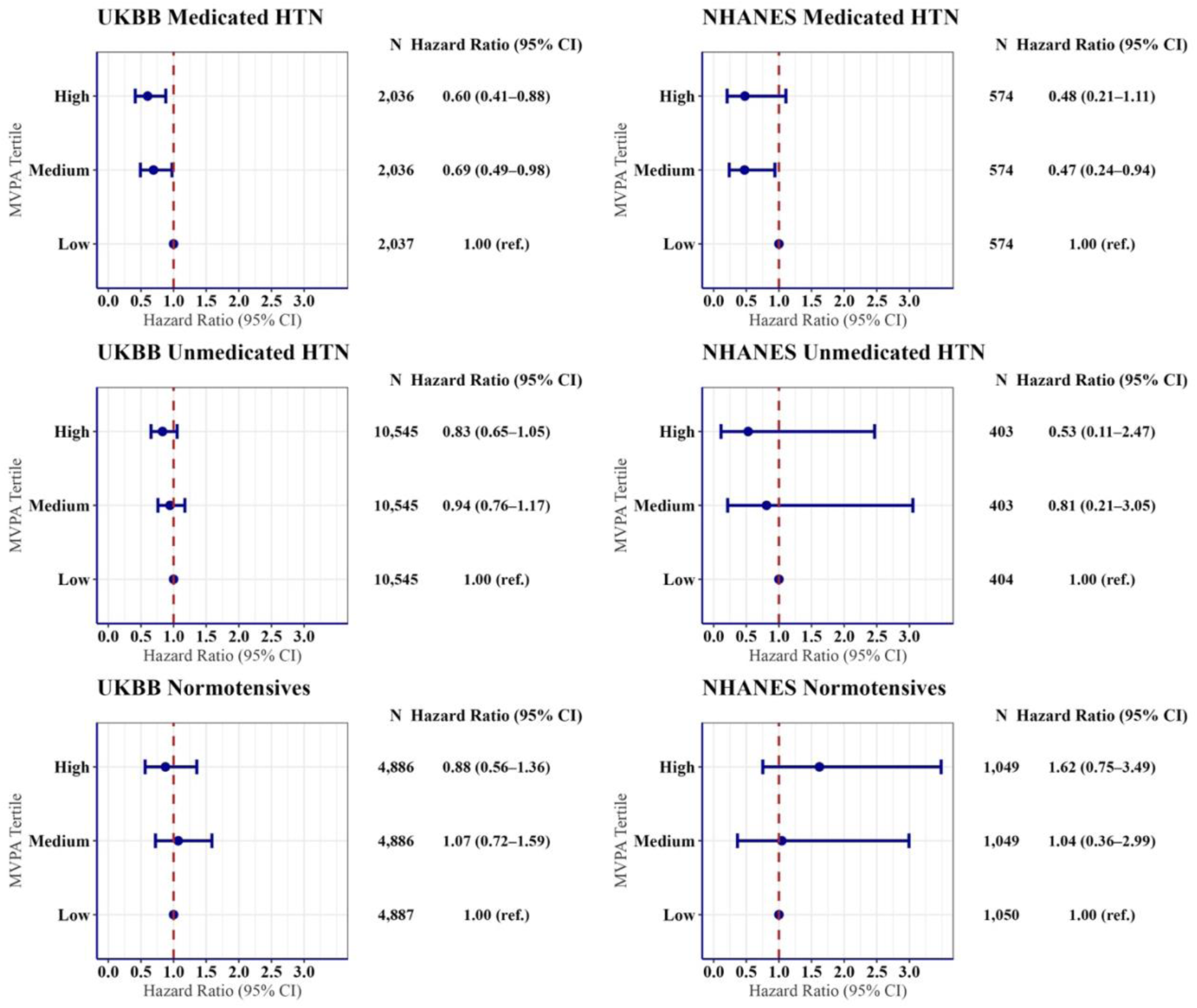
Hazard ratios for all-cause mortality across moderate-to-vigorous physical activity tertiles within hypertension subgroups in UK Biobank and NHANES *Each subgroup was stratified into MVPA tertiles (low, medium, high), with the low MVPA tertile serving as the reference category (HR = 1.00). HRs are presented with 95% confidence intervals. Models adjusted for age, sex, smoking status, fruit and vegetables intake, previous CVD, family history of CVD, sleep duration, educational attainment, alcohol consumption, ethnicity, and light physical activity energy expenditure*.

**Supplementary Figure 5:**
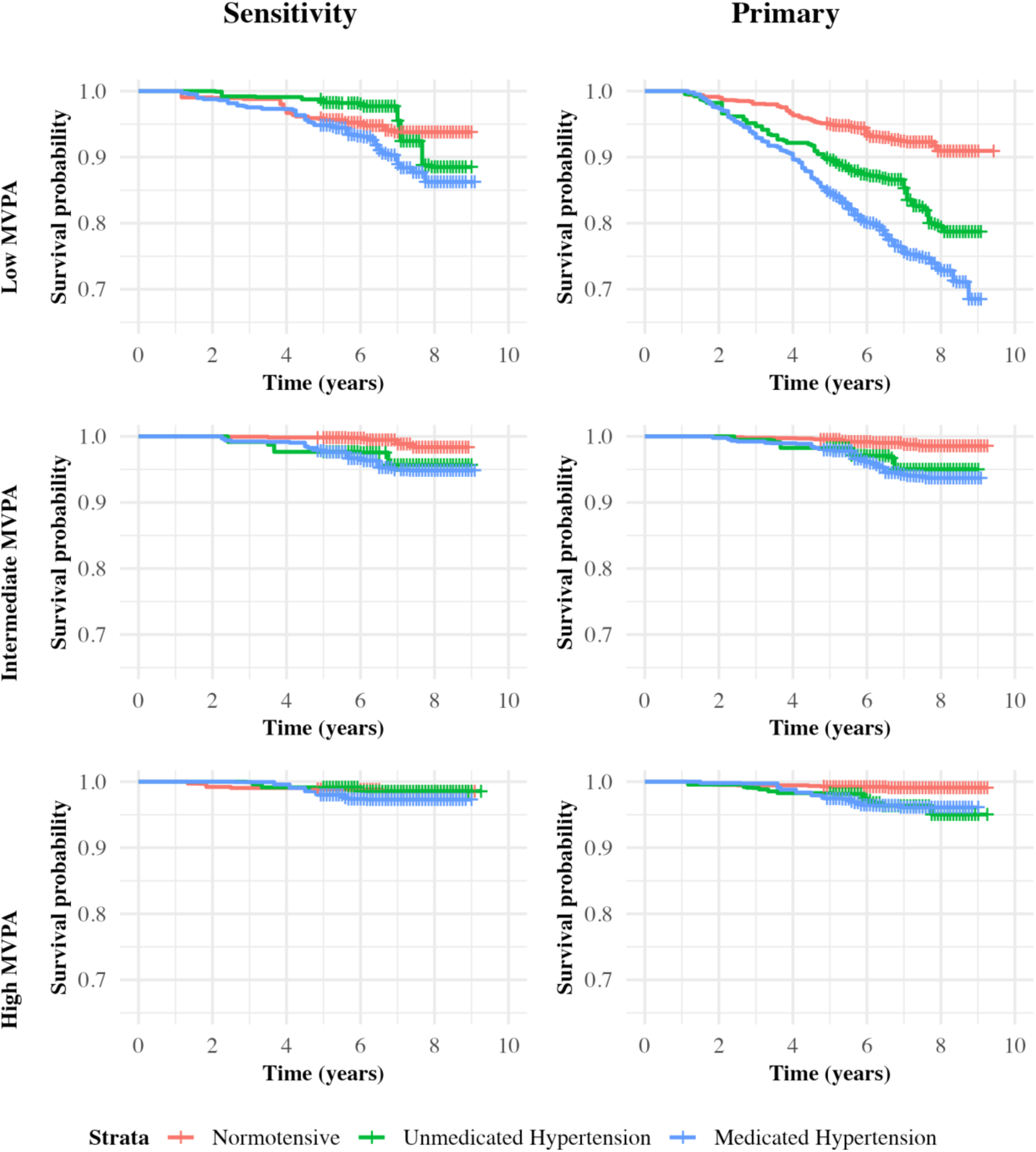
Weighted Kaplan–Meier survival curves for all-cause mortality by hypertension status stratified by moderate-to-vigorous physical activity tertiles: primary analysis and sensitivity analysis restricted to adults aged 40–69 in NHANES *The primary analysis includes all eligible NHANES participants aged ≥20 years. The sensitivity analysis is restricted to adults aged 40–69 years, to align with the age range of participants in UKBB*.

**Supplementary Figure 6:**
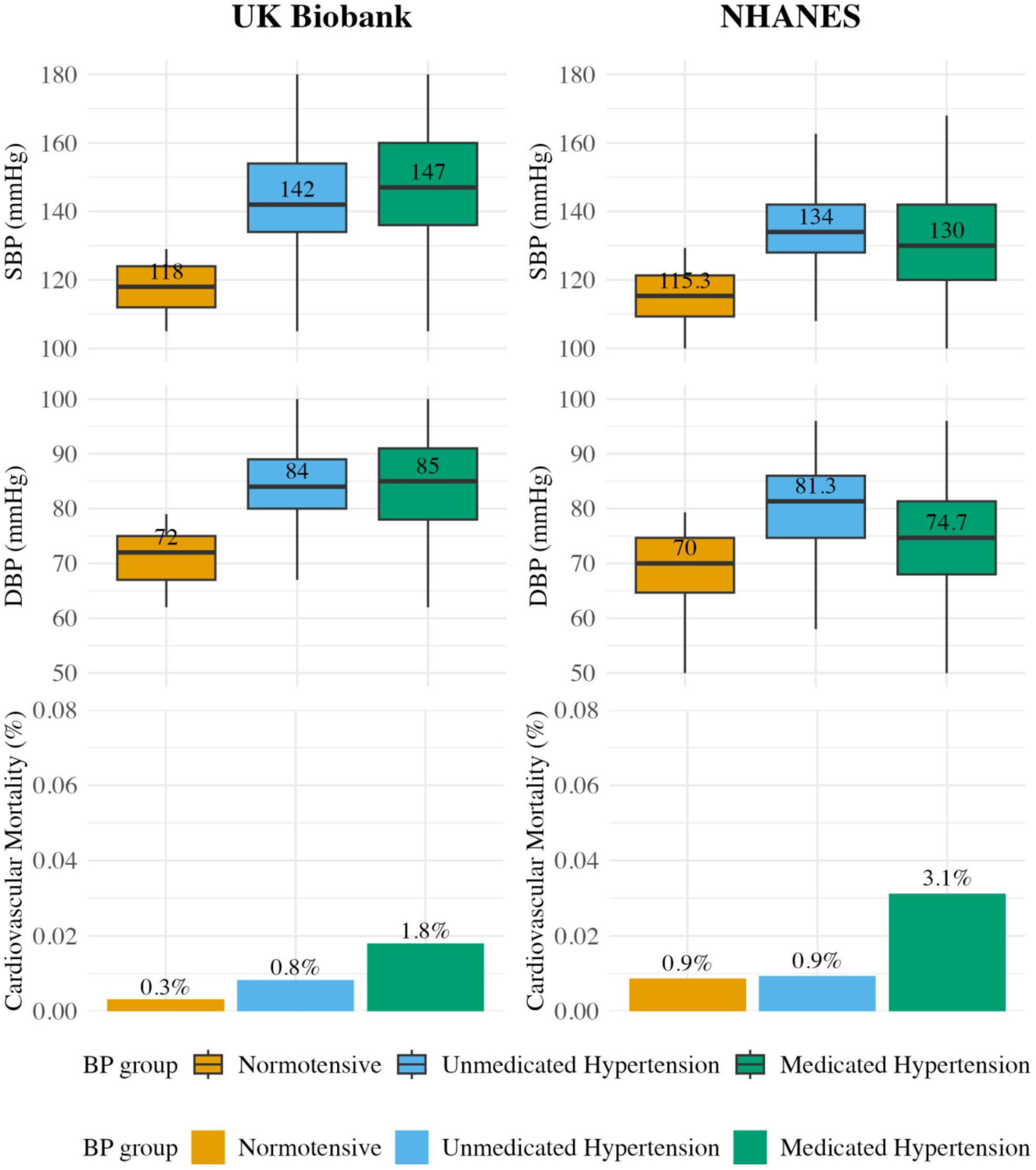
Weighted distribution of blood pressure and cardiovascular mortality by hypertension groups in UK Biobank and NHANES: sensitivity analysis restricted to adults aged 40– 69 years in NHANES *The primary analysis includes all eligible NHANES participants aged ≥20 years. The sensitivity analysis is restricted to adults aged 40–69 years, to align with the age range of participants in UKBB*.

**Supplementary Figure 7:**
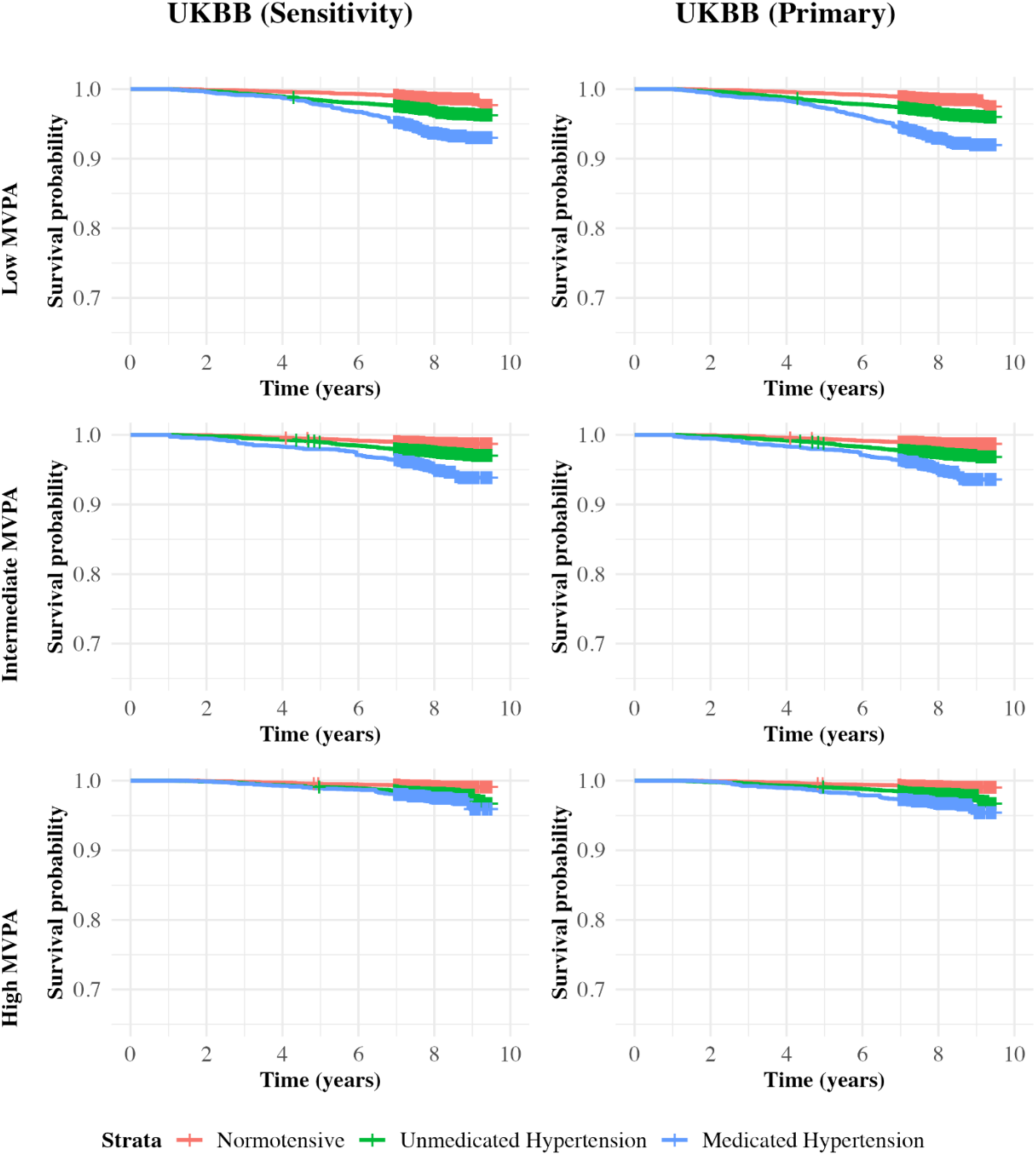
Weighted Kaplan–Meier survival curves for all-cause mortality by hypertension status stratified by moderate-to-vigorous physical activity tertiles in UK Biobank: primary analysis and sensitivity analysis excluding participants with prior cardiovascular disease

**Supplementary Figure 8:**
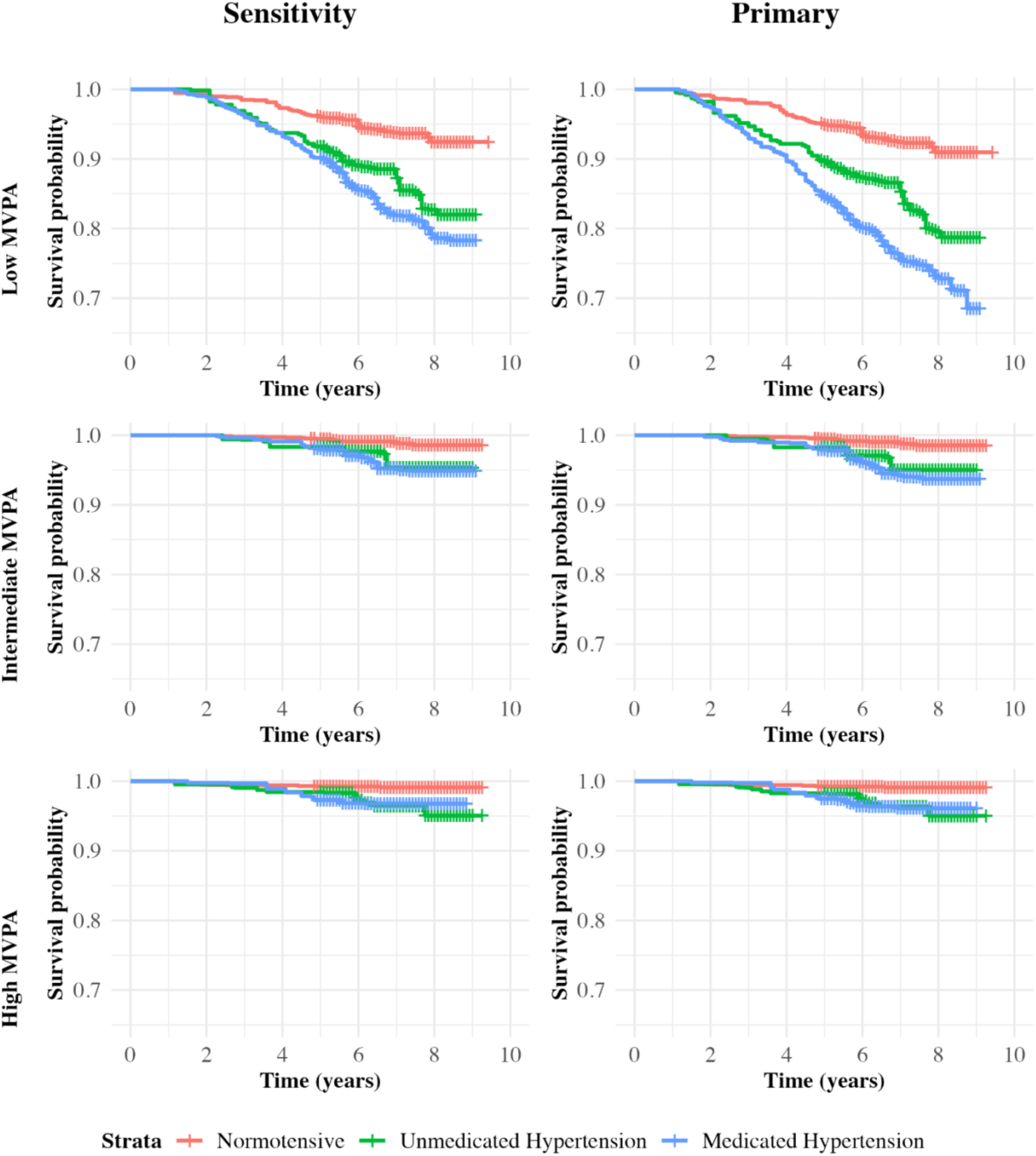
Weighted Kaplan–Meier survival curves for all-cause mortality by hypertension status stratified by moderate-to-vigorous physical activity tertiles: primary and sensitivity analyses excluding participants with prior cardiovascular disease in NHANES

**Supplementary Figure 9:**
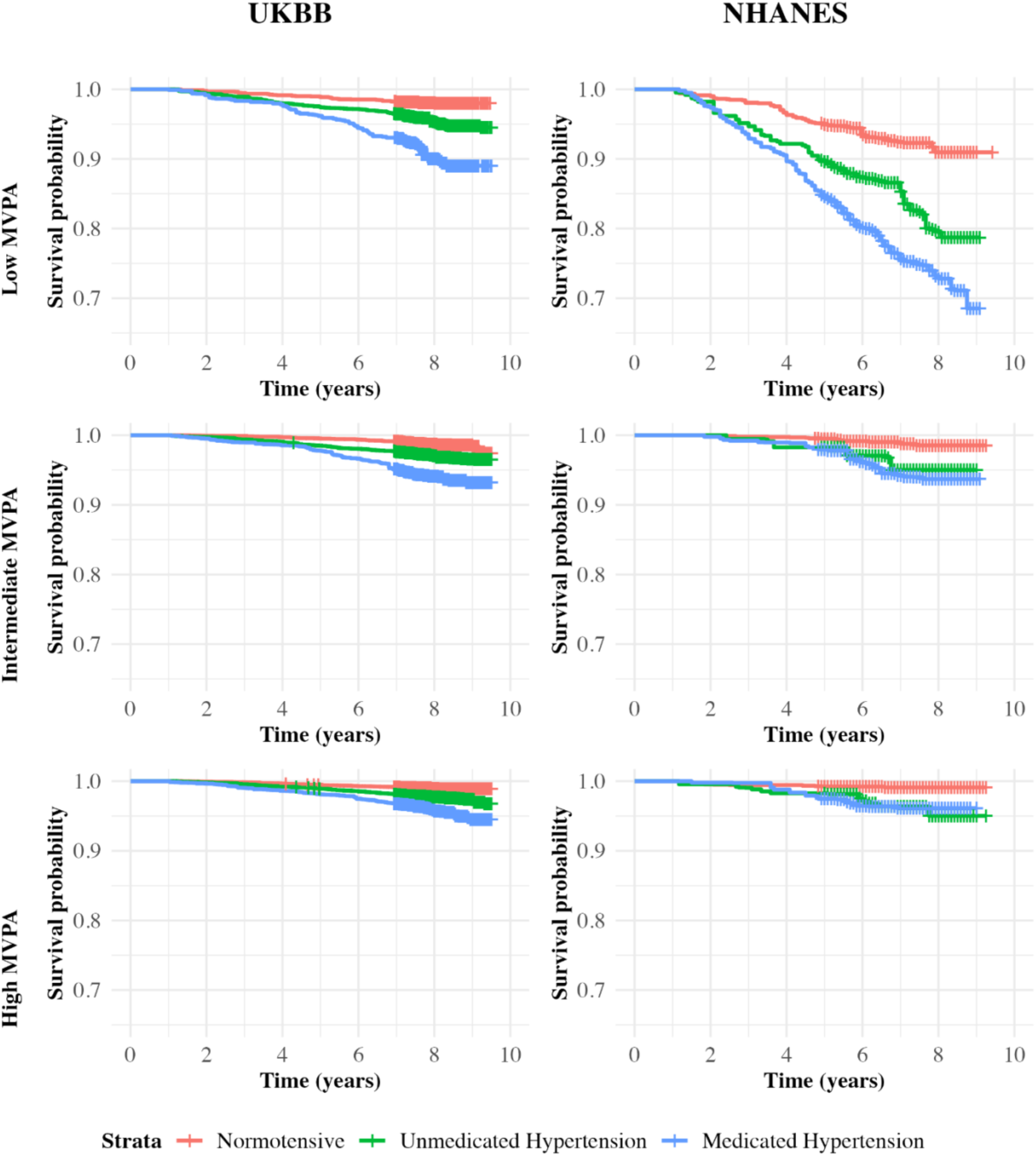
Weighted Kaplan–Meier survival curves for all-cause mortality by hypertension status stratified by NHANES-specific moderate-to-vigorous physical activity tertiles in UK Biobank and NHANES *MVPA tertiles were defined using the NHANES distribution as follows: Low (0.29–10.72 min/day), Intermediate (10.73–24.78 min/day), and High (≥24.79 min/day). These thresholds were applied uniformly to both UKBB and NHANES*.

**Supplementary Figure 10:**
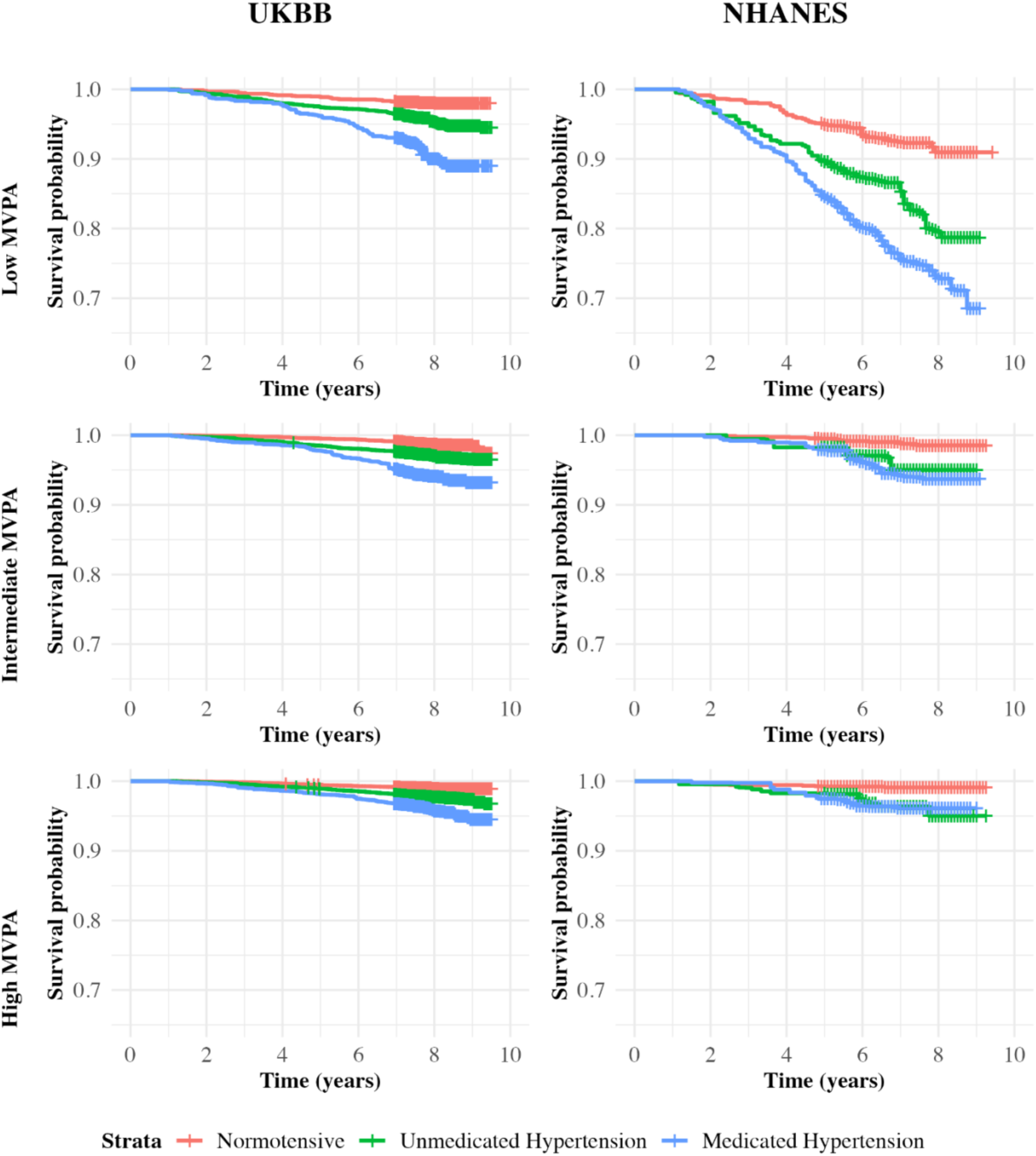
Unweighted Kaplan-Meier survival curves for all-cause mortality by hypertension groups stratified by moderate-to-vigorous physical activity tertiles in UK Biobank and NHANES *The unweighted sensitivity analysis includes all eligible participants aged ≥20 years in both cohorts and was conducted to assess whether Kaplan-Meier survival curves were influenced by weighting approaches*.

**Supplementary Table 1:** Weighted baseline demographic, behavioural, and clinical characteristics of participants stratified by hypertension status in UK Biobank and NHANES

|  | *Overall*<br>*UKBB* | *Normotensive*<br>*UKBB* | *Unmedicated*<br>*HTN UKBB* | *Medicated*<br>*HTN*<br>*UKBB* | *Overall*<br>*NHANES* | *Normotensive*<br>*NHANES* | *Unmedicated*<br>*HTN*<br>*NHANES* | *Medicated*<br>*HTN*<br>*NHANES* |
| --- | --- | --- | --- | --- | --- | --- | --- | --- |
| *n* | 53049 | 15750 | 31110 | 6190 | 157714415 | 79197815 | 30617498 | 47899102 |
| *Age, mean (s.d.),*<br>*years* | 58.24<br>(8.26) | 55.31 (7.42) | 58.58 (8.18) | 63.93<br>(7.29) | 49.31<br>(16.58) | 41.93 (14.71) | 50.50<br>(15.28) | 60.76<br>(13.26) |
| *Female, n (%)* | 27888<br>(52.6) | 10446 (66.3) | 14668 (47.1) | 2774<br>(44.8) | 81057233<br>(51.4) | 42524713<br>(53.7) | 12576778<br>(41.1) | 25955742<br>(54.2) |
| *Ethnicity, n (%)* |  |  |  |  |  |  |  |  |
| *White* | 50146<br>(94.5) | 14800 (94.0) | 29504 (94.8) | 5841<br>(94.4) | 107949339<br>(68.4) | 52628154<br>(66.5) | 20917441<br>(68.3) | 34403744<br>(71.8) |
| *Black* | 887<br>(1.7) | 251 (1.6) | 488 (1.6) | 148 (2.4) | 16984269<br>(10.8) | 6604445<br>(8.3) | 3604798<br>(11.8) | 6775026<br>(14.1) |
| *Asian* | 1301<br>(2.5) | 420 (2.7) | 743 (2.4) | 138 (2.2) | 6926901<br>(4.4) | 4221495<br>(5.3) | 1249162<br>(4.1) | 1456244<br>(3.0) |
| *Other* | 716<br>(1.3) | 278 (1.8) | 375 (1.2) | 63 (1.0) | 25853906<br>(16.4) | 15743722<br>(19.9) | 4846097<br>(15.8) | 5264087<br>(11.0) |
| *Smoking*<br>*history, n (%)* |  |  |  |  |  |  |  |  |
| *Never* | 30745<br>(58.0) | 9532 (60.5) | 18014 (57.9) | 3200<br>(51.7) | 80105586<br>(56.6) | 46142249<br>(61.4) | 15725290<br>(55.5) | 18238046<br>(48.1) |
| *Previous* | 17862<br>(33.7) | 4768 (30.3) | 10522 (33.8) | 2572<br>(41.6) | 35028853<br>(24.8) | 14303349<br>(19.0) | 7317600<br>(25.8) | 13407905<br>(35.4) |
| *Current* | 4442<br>(8.4) | 1450 (9.2) | 2575 (8.3) | 418 (6.7) | 26302250<br>(18.6) | 14763827<br>(19.6) | 5305744<br>(18.7) | 6232680<br>(16.5) |
| *Alcohol daily*<br>*consumption,*<br>*mean (s.d.)* | 1.93<br>(2.25) | 1.53 (1.71) | 2.08 (2.37) | 2.18<br>(2.65) | 2.57 (2.23) | 2.61 (2.21) | 2.64 (2.22) | 2.44 (2.27) |
| *Education, n (%)* |  |  |  |  |  |  |  |  |
| *High school*<br>*graduate, GED*<br>*or equivalent* | 13702.4<br>(25.8) | 4116.0 (26.1) | 8068.1<br>(25.9) | 1518.3<br>(24.5) | 33127235.1<br>(21.0) | 14907871.6<br>(18.8) | 7270395.3<br>(23.7) | 10948968.3<br>(22.9) |
| *Some college or*<br>*AA degree* | 10085.2<br>(19.0) | 2627.2 (16.7) | 6199.9<br>(19.9) | 1258.2<br>(20.3) | 50793895.5<br>(32.2) | 25179272.6<br>(31.8) | 9237213.9<br>(30.2) | 16377409.0<br>(34.2) |
| *College*<br>*graduate* | 22327.1<br>(42.1) | 7590.5 (48.2) | 12688.6<br>(40.8) | 2048.0<br>(33.1) | 49859228.0<br>(31.6) | 28494089.8<br>(36.0) | 9656906.7<br>(31.5) | 11708231.4<br>(24.5) |
| *Other* | 6934.4<br>(13.1) | 1416.2 (9.0) | 4153.2<br>(13.4) | 1365.0<br>(22.1) | 23876102.5<br>(15.1) | 10592293.8<br>(13.4) | 4448313.5<br>(14.5) | 8835495.3<br>(18.5) |
| *Fruit and*<br>*vegetable*<br>*consumption,* | 7.77<br>(4.48) | 7.83 (4.65) | 7.68 (4.36) | 8.07<br>(4.67) | 4.76 (3.53) | 4.84 (3.86) | 4.67 (3.06) | 4.68 (3.22) |
| *mean (s.d.),*<br>*servings/day* |  |  |  |  |  |  |  |  |
| *Previous CVD,*<br>*n(%)* | 3729.8<br>(7.0) | 688.1 (4.4) | 2123.7 (6.8) | 918.0<br>(14.8) | 14562762.3<br>(9.2) | 2867626.7<br>(3.6) | 1538512.5<br>(5.0) | 10156623.1<br>(21.3) |
| *Family history*<br>*of CVD, n (%)* | 26826.5<br>(50.6) | 7057.1 (44.8) | 15907.7<br>(51.1) | 3861.7<br>(62.4) | 19830689.8<br>(12.8) | 7972862.9<br>(10.3) | 3208760.6<br>(10.7) | 8649066.3<br>(18.4) |
| *Non-exerciser,*<br>*n (%)* | 17107.5<br>(32.2) | 4303.3 (27.3) | 10158.2<br>(32.7) | 2645.9<br>(42.7) | 73218637.2<br>(46.4) | 31048363.9<br>(39.2) | 14762375.2<br>(48.2) | 27407898.1<br>(57.2) |
| *Sleep, mean*<br>*(s.d.),*<br>*minutes/day* | 440.08<br>(91.78) | 441.69<br>(96.80) | 439.74<br>(90.59) | 437.69<br>(84.22) | 390.67<br>(125.14) | 402.08<br>(119.73) | 380.12<br>(127.52) | 378.54<br>(130.58) |
| *HbA1C, mean*<br>*(s.d.), %* | 5.35<br>(0.51) | 5.25 (0.38) | 5.36 (0.52) | 5.57<br>(0.67) | 5.67 (0.98) | 5.44 (0.73) | 5.70 (1.04) | 6.03 (1.18) |
| *HDL, mean*<br>*(s.d.), mg/dL* | 56.44<br>(14.45) | 58.57 (14.36) | 56.01<br>(14.37) | 53.18<br>(14.29) | 52.86<br>(15.82) | 53.63 (15.14) | 52.87<br>(18.13) | 51.56<br>(15.27) |
| *LDL, mean*<br>*(s.d.), mg/dL* | 136.09<br>(32.18) | 130.42<br>(30.07) | 140.58<br>(32.09) | 127.91<br>(33.87) | 114.61<br>(35.50) | 114.51<br>(33.06) | 119.88<br>(36.35) | 112.18<br>(38.43) |
| *triglyceride,*<br>*mean (s.d.),*<br>*mg/dL* | 145.72<br>(87.33) | 121.32<br>(70.88) | 153.78<br>(91.03) | 167.36<br>(92.81) | 129.18<br>(116.30) | 115.45<br>(128.78) | 136.83<br>(92.45) | 147.04<br>(102.59) |
| *SBP, mean*<br>*(s.d.), mmHg* | 136.02<br>(17.92) | 117.29 (7.33) | 143.28<br>(14.45) | 147.20<br>(16.88) | 122.23<br>(16.00) | 112.54 (8.74) | 133.80<br>(12.42) | 130.84<br>(17.41) |
| *DBP, mean*<br>*(s.d.), mmHg* | 81.26<br>(10.12) | 71.23 (5.06) | 85.42 (8.41) | 85.84<br>(9.68) | 70.67<br>(10.76) | 67.20 (7.87) | 79.07<br>(10.03) | 71.03<br>(12.23) |
| *Waist*<br>*circumference,*<br>*mean (s.d.), cm* | 88.27<br>(13.09) | 82.34 (11.14) | 89.76<br>(12.65) | 95.87<br>(13.81) | 100.20<br>(16.11) | 95.60 (14.58) | 102.14<br>(16.01) | 106.81<br>(16.12) |
| *Body Mass*<br>*Index, mean*<br>*(s.d.), kg/m<sup>2</sup>* | 26.70<br>(4.59) | 24.88 (3.69) | 27.11 (4.49) | 29.28<br>(5.35) | 29.25<br>(6.82) | 27.75 (5.92) | 29.77 (6.92) | 31.43<br>(7.49) |
| *CVD*<br>*mortality, n (%)* | 328.2<br>(0.7) | 36.8 (0.2) | 199.7 (0.7) | 91.7 (1.7) | 3634417.5<br>(2.3) | 659032.3<br>(0.8) | 515765.1<br>(1.7) | 2459620.1<br>(5.1) |
| *Daily MVPA*<br>*duration, mean*<br>*(s.d.),*<br>*minutes/day* | 40.01<br>(25.23) | 42.47 (25.31) | 40.09<br>(25.24) | 33.36<br>(23.76) | 21.04<br>(17.37) | 24.69 (17.38) | 22.90<br>(17.28) | 13.82<br>(15.05) |
| **Abbreviations:**<br>*AA, Associate's degree;*<br>*GED, General Educational Development;*<br>*HDL, high-density lipoprotein cholesterol;*<br>*LDL, low-density lipoprotein cholesterol;* |  |  |  |  |  |  |  |  |

**Supplementary Table 2:** Unweighted baseline demographic, behavioural, and clinical characteristics of participants stratified by hypertension status in UK Biobank and NHANES

|  | *Overall*<br>*UKBB* | *Normotensive*<br>*UKBB* | *Unmedicated*<br>*HTN UKBB* | *Medicated*<br>*HTN*<br>*UKBB* | *Overall*<br>*NHANES* | *Normotensive*<br>*NHANES* | *Unmedicated*<br>*HTN*<br>*NHANES* | *Medicated*<br>*HTN*<br>*NHANES* |
| --- | --- | --- | --- | --- | --- | --- | --- | --- |
| *n* | 63452 | 16814 | 37747 | 8891 | 7397 | 3445 | 1414 | 2538 |
| *Age, mean (s.d.),*<br>*years* | 61.57 (7.86) | 58.69 (7.80) | 61.88 (7.72) | 65.72<br>(6.33) | 50.72<br>(17.28) | 41.89 (15.26) | 51.76<br>(15.88) | 62.12<br>(13.31) |
| *Female, n (%)* | 35790 (56.4) | 11783 (70.1) | 19850 (52.6) | 4157<br>(46.8) | 3749<br>(50.7) | 1837 (53.3) | 560 (39.6) | 1352<br>(53.3) |
| *Ethnicity, n (%)* |  |  |  |  |  |  |  |  |
| *White* | 61597 (97.1) | 16255 (96.7) | 36741 (97.3) | 8601<br>(96.7) | 3118<br>(42.2) | 1434 (41.6) | 577 (40.8) | 1107<br>(43.6) |
| *Black* | 514 (0.8) | 128 (0.8) | 275 (0.7) | 111 (1.2) | 1719<br>(23.2) | 584 (17.0) | 360 (25.5) | 775 (30.5) |
| *Asian* | 675 (1.1) | 203 (1.2) | 374 (1.0) | 98 (1.1) | 796<br>(10.8) | 472 (13.7) | 144 (10.2) | 180 (7.1) |
| *Other* | 666 (1.0) | 228 (1.4) | 357 (0.9) | 81 (0.9) | 1764<br>(23.8) | 955 (27.7) | 333 (23.6) | 476 (18.8) |
| *Smoking*<br>*history, n (%)* |  |  |  |  |  |  |  |  |
| *Never* | 36444 (57.4) | 10133 (60.3) | 21709 (57.5) | 4602<br>(51.8) | 3685<br>(57.2) | 1999 (61.7) | 721 (56.0) | 965 (50.3) |
| *Previous* | 22795 (35.9) | 5424 (32.3) | 13563 (35.9) | 3808<br>(42.8) | 1503<br>(23.3) | 582 (18.0) | 294 (22.8) | 627 (32.7) |
| *Current* | 4213 (6.6) | 1257 (7.5) | 2475 (6.6) | 481 (5.4) | 1259<br>(19.5) | 660 (20.4) | 273 (21.2) | 326 (17.0) |
| *Alcohol daily*<br>*consumption,*<br>*mean (s.d.)* | 1.97 (2.17) | 1.58 (1.70) | 2.08 (2.25) | 2.22<br>(2.51) | 2.62<br>(2.33) | 2.70 (2.39) | 2.72 (2.29) | 2.41<br>(2.26) |
| *Education, n (%)* |  |  |  |  |  |  |  |  |
| *High school*<br>*graduate, GED*<br>*or equivalent* | 15615 (24.6) | 3898 (23.2) | 9430 (25.0) | 2287<br>(25.7) | 1617<br>(21.9) | 663 (19.3) | 341 (24.1) | 613 (24.2) |
| *Some college or*<br>*AA degree* | 11759 (18.5) | 2877 (17.1) | 7205 (19.1) | 1677<br>(18.9) | 2252<br>(30.5) | 1067 (31.0) | 411 (29.1) | 774 (30.5) |
| *College*<br>*graduate* | 28325 (44.6) | 8592 (51.1) | 16429 (43.5) | 3304<br>(37.2) | 1921<br>(26.0) | 1077 (31.3) | 365 (25.8) | 479 (18.9) |
| *Other* | 7753 (12.2) | 1447 (8.6) | 4683 (12.4) | 1623<br>(18.3) | 1602<br>(21.7) | 636 (18.5) | 296 (20.9) | 670 (26.4) |
| *Fruit and*<br>*vegetable*<br>*consumption,* | 8.04 (4.35) | 8.13 (4.46) | 7.96 (4.29) | 8.19<br>(4.40) | 4.61<br>(3.36) | 4.71 (3.53) | 4.60 (3.24) | 4.48<br>(3.17) |
| *mean (s.d.),*<br>*servings/day* |  |  |  |  |  |  |  |  |
| *Previous*<br>*CVD, n (%)* | 5199 (8.2) | 903 (5.4) | 2923 (7.7) | 1373<br>(15.4) | 827<br>(11.2) | 138 (4.0) | 80 (5.7) | 609 (24.2) |
| *Family history*<br>*of CVD, n (%)* | 34776 (54.8) | 8235 (49.0) | 20816 (55.1) | 5725<br>(64.4) | 837<br>(11.6) | 316 (9.4) | 139 (10.1) | 382 (15.4) |
| *Non-exerciser,*<br>*n (%)* | 20095 (31.7) | 4545 (27.0) | 12080 (32.0) | 3470<br>(39.0) | 3744<br>(50.6) | 1493 (43.3) | 717 (50.7) | 1534<br>(60.4) |
| *Sleep, mean*<br>*(s.d.),*<br>*minutes/day* | 442.38<br>(90.04) | 444.76<br>(92.59) | 442.10<br>(90.37) | 439.09<br>(83.33) | 380.91<br>(124.45) | 394.35<br>(119.28) | 371.41<br>(124.65) | 367.96<br>(129.31) |
| *HbA1C, mean*<br>*(s.d.), %* | 5.38 (0.50) | 5.28 (0.37) | 5.37 (0.48) | 5.58<br>(0.67) | 5.80<br>(1.14) | 5.52 (0.88) | 5.82 (1.20) | 6.18<br>(1.30) |
| *HDL, mean*<br>*(s.d.), mg/dL* | 57.67 (14.84) | 60.09 (14.66) | 57.50<br>(14.79) | 53.87<br>(14.51) | 52.47<br>(15.53) | 53.05 (14.83) | 52.50<br>(17.17) | 51.65<br>(15.47) |
| *LDL, mean*<br>*(s.d.), mg/dL* | 138.08<br>(32.60) | 133.62<br>(30.93) | 142.46<br>(32.28) | 127.94<br>(33.57) | 113.20<br>(35.47) | 114.08<br>(34.05) | 118.57<br>(35.46) | 109.51<br>(37.02) |
| *triglyceride,*<br>*mean (s.d.),*<br>*mg/dL* | 145.93<br>(84.77) | 122.16<br>(69.78) | 152.11<br>(87.49) | 164.66<br>(89.40) | 127.58<br>(120.99) | 117.59<br>(137.19) | 134.05<br>(99.60) | 138.44<br>(103.67) |
| *SBP, mean*<br>*(s.d.), mmHg* | 138.04<br>(18.32) | 117.65 (7.25) | 144.81<br>(14.81) | 147.89<br>(16.93) | 123.87<br>(16.98) | 112.58 (8.75) | 135.73<br>(12.87) | 132.58<br>(17.80) |
| *DBP, mean*<br>*(s.d.), mmHg* | 81.53 (10.07) | 71.20 (5.05) | 85.20 (8.52) | 85.45<br>(9.62) | 70.37<br>(11.10) | 66.84 (8.03) | 78.21<br>(10.84) | 70.78<br>(12.44) |
| *Waist*<br>*circumference,*<br>*mean (s.d.), cm* | 88.11 (12.95) | 82.10 (11.00) | 89.16<br>(12.49) | 95.05<br>(13.54) | 99.70<br>(16.35) | 94.69 (14.81) | 101.23<br>(16.02) | 105.88<br>(16.32) |
| *Body mass*<br>*index, mean*<br>*(s.d.), kg/m<sup>2</sup>* | 26.63 (4.49) | 24.75 (3.57) | 26.93 (4.37) | 28.91<br>(5.13) | 29.20<br>(6.97) | 27.57 (6.03) | 29.57 (7.01) | 31.24<br>(7.57) |
| *CVD*<br>*mortality, n (%)* | 478 (0.8) | 50 (0.3) | 292 (0.8) | 136 (1.8) | 233 (3.1) | 40 (1.2) | 27 (1.9) | 166 (6.5) |
| *Daily MVPA*<br>*duration, mean*<br>*(s.d.),*<br>*minutes/day* | 38.41 (24.76) | 41.03 (25.08) | 38.70<br>(24.83) | 32.23<br>(22.76) | 19.75<br>(17.37) | 24.17 (17.50) | 22.13<br>(17.88) | 12.42<br>(14.22) |
| **Abbreviations:**<br>*AA*, Associate's degree;<br>*GED*, General Educational Development;<br>*HDL*, high-density lipoprotein cholesterol;<br>*LDL*, low-density lipoprotein cholesterol; |  |  |  |  |  |  |  |  |

**Supplementary Table 3:** Number of all-cause mortality events by moderate-to-vigorous physical activity tertiles within hypertension subgroups in UK Biobank and NHANES

|  | *UKBB*<br>*medicated HTN* | *UKBB*<br>*unmedicated*<br>*HTN* | *UKBB*<br>*normotensive* | *NHANES*<br>*medicated HTN* | *NHANES*<br>*unmedicated*<br>*HTN* | *NHANES*<br>*normotensive* |
| --- | --- | --- | --- | --- | --- | --- |
| *Overall* | 367 | 1,032 | 259 | 260 | 78 | 78 |
| *MVPA Tertiles* |  |  |  |  |  |  |
| *Low* | 166 | 432 | 116 | 181 | 56 | 54 |
| *Intermediate* | 105 | 337 | 83 | 53 | 13 | 12 |
| *High* | 96 | 263 | 60 | 26 | 9 | 12 |
| *Values represent the total number of all-cause mortality events observed during follow-up in each MVPA tertile, stratified by hypertension status.* |  |  |  |  |  |  |

## References

1. Lim SS, Vos T, Flaxman AD, et al. A comparative risk assessment of burden of disease and injury attributable to 67 risk factors and risk factor clusters in 21 regions, 1990-2010: a systematic analysis for the Global Burden of Disease Study 2010. Lancet 2012; 380: 2224–2260. 2012/12/19. DOI: 10.1016/s0140-6736(12)61766-8.

2. Forouzanfar MH, Liu P, Roth GA, et al. Global Burden of Hypertension and Systolic Blood Pressure of at Least 110 to 115 mm Hg, 1990-2015. Jama 2017; 317: 165–182. 2017/01/18. DOI: 10.1001/jama.2016.19043.

3. Mills KT, Stefanescu A and He J. The global epidemiology of hypertension. Nat Rev Nephrol 2020; 16: 223–237. 2020/02/07. DOI: 10.1038/s41581-019-0244-2.

4. Lewington S, Clarke R, Qizilbash N, et al. Age-specific relevance of usual blood pressure to vascular mortality: a meta-analysis of individual data for one million adults in 61 prospective studies. Lancet 2002; 360: 1903–1913. 2002/12/21. DOI: 10.1016/s0140-6736(02)11911-8.

5. Whelton PK, Carey RM, Aronow WS, et al. 2017 ACC/AHA/AAPA/ABC/ACPM/AGS/APhA/ASH/ASPC/NMA/PCNA guideline for the prevention, detection, evaluation, and management of high blood pressure in adults: a report of the American College of Cardiology/American Heart Association Task Force on Clinical Practice Guidelines. Journal of the American College of Cardiology 2018; 71: e127–e248.

6. National Institute for Health and Care Excellence: Guidelines. Hypertension in adults: diagnosis and management. London: National Institute for Health and Care Excellence (NICE) Copyright © NICE 2023., 2023.

7. Li J, Lei L, Li Y, et al. Effect of Intensive Blood Pressure Control on Stroke: A Prespecified Secondary Analysis of the ESPRIT Trial. JACC 2025.

8. Lesniak KT and Dubbert PM. Exercise and hypertension. Curr Opin Cardiol 2001; 16: 356–359. 2001/11/13. DOI: 10.1097/00001573-200111000-00007.

9. Hou X, Yue S, Xu Z, et al. Joint Modifiable Risk Factor Control and Incident Stroke in Hypertensive Patients. J Clin Hypertens (Greenwich) 2024; 26: 1274–1283. 2024/09/28 22:42. DOI: 10.1111/jch.14905.

10. Maris SA, Meyer KM, Murray G, et al. Physical Activity and the Acute Hemodynamic Response to ACE Inhibition in Hypertension. American Journal of Lifestyle Medicine 2022; 16: 538–545. DOI: 10.1177/1559827620935367.

11. Goessler K, Polito M and Cornelissen VA. Effect of exercise training on the renin-angiotensin-aldosterone system in healthy individuals: a systematic review and meta-analysis. Hypertens Res 2016; 39: 119–126. 2015/09/25. DOI: 10.1038/hr.2015.100.

12. Kapteyn A, Banks J, Hamer M, et al. What they say and what they do: comparing physical activity across the USA, England and the Netherlands. J Epidemiol Community Health 2018; 72: 471–476. 2018/04/13. DOI: 10.1136/jech-2017-209703.

13. Acosta E, Mehta N, Myrskylä M, et al. Cardiovascular Mortality Gap Between the United States and Other High Life Expectancy Countries in 2000-2016. J Gerontol B Psychol Sci Soc Sci 2022; 77: S148–s157. 2022/02/24. DOI: 10.1093/geronb/gbac032.

14. Pritchard C, Porter S, Rosenorn-Lanng E, et al. Mortality in the USA, the UK and Other Western Countries, 1989–2015: What Is Wrong With the US? International Journal of Health Services 2021; 51: 59–66. DOI: 10.1177/0020731420965130.

15. Allen N, Sudlow C, Downey P, et al. UK Biobank: Current status and what it means for epidemiology. Health Policy and Technology 2012; 1: 123–126.

16. Sudlow C, Gallacher J, Allen N, et al. UK biobank: an open access resource for identifying the causes of a wide range of complex diseases of middle and old age. PLoS Med 2015; 12: e1001779. 20150331. DOI: 10.1371/journal.pmed.1001779.

17. Statistics NCfH. NHANES Survey Methods and Analytic Guidelines. 2024.

18. Terry AL, Chiappa MM, McAllister J, et al. Plan and Operations of the National Health and Nutrition Examination Survey, August 2021-August 2023. Vital and Health statistics Ser 1, Programs and Collection Procedures 2024: 1–21.

19. Fry A, Littlejohns TJ, Sudlow C, et al. Comparison of Sociodemographic and Health-Related Characteristics of UK Biobank Participants With Those of the General Population. Am J Epidemiol 2017; 186: 1026–1034. DOI: 10.1093/aje/kwx246.

20. Biobank U. Mortality data: linkage to death registries. 2023.

21. Doherty A, Jackson D, Hammerla N, et al. Large scale population assessment of physical activity using wrist worn accelerometers: the UK biobank study. PloS one 2017; 12: e0169649.

22. Prevention CfDCa. NCHS Data Linked to NDI Mortality Files, https://www.cdc.gov/nchs/data-linkage/mortality.htm (2024, accessed June 25th 2025).

23. Koffman L and Muschelli J. Minute level step counts and physical activity data from the National Health and Nutrition Examination Survey (NHANES) 2011-2014.

24. Stamatakis E, Ahmadi MN, Gill JMR, et al. Association of wearable device-measured vigorous intermittent lifestyle physical activity with mortality. Nat Med 2022; 28: 2521–2529. 20221208. DOI: 10.1038/s41591-022-02100-x.

25. Pavey TG, Gilson ND, Gomersall SR, et al. Field evaluation of a random forest activity classifier for wrist-worn accelerometer data. J Sci Med Sport 2017; 20: 75–80. 20160623. DOI: 10.1016/j.jsams.2016.06.003.

26. Ahmadi MN, Clare PJ, Katzmarzyk PT, et al. Vigorous physical activity, incident heart disease, and cancer: how little is enough? Eur Heart J 2022; 43: 4801–4814. DOI: 10.1093/eurheartj/ehac572.

27. Stamatakis E, Ahmadi MN, Friedenreich CM, et al. Vigorous Intermittent Lifestyle Physical Activity and Cancer Incidence Among Nonexercising Adults: The UK Biobank Accelerometry Study. JAMA Oncol 2023; 9: 1255–1259. DOI: 10.1001/jamaoncol.2023.1830.

28. Koemel NA, Ahmadi MN, Biswas RK, et al. Vigorous intermittent lifestyle physical activity (VILPA) and mortality risk among US adults: a wearables-based national cohort study. Int J Behav Nutr Phys Act 2026; 23 20260130. DOI: 10.1186/s12966-026-01876-2.

29. Stamatakis E, Koemel NA, Biswas RK, et al. Minimum and optimal combined variations in sleep, physical activity, and nutrition in relation to all-cause mortality risk. BMC Med 2025; 23: 111. 20250226. DOI: 10.1186/s12916-024-03833-x.

30. Koemel NA, Ahmadi MN, Biswas RK, et al. Can incidental physical activity offset the deleterious associations of sedentary behaviour with major adverse cardiovascular events? Eur J Prev Cardiol 2025; 32: 77–85. DOI: 10.1093/eurjpc/zwae316.

31. Hildebrand M, VT VANH, Hansen BH, et al. Age group comparability of raw accelerometer output from wrist- and hip-worn monitors. Med Sci Sports Exerc 2014; 46: 1816–1824. DOI: 10.1249/MSS.0000000000000289.

32. Vidal-Petiot E. Thresholds for Hypertension Definition, Treatment Initiation, and Treatment Targets: Recent Guidelines at a Glance. Circulation 2022; 146: 805–807. DOI: doi:10.1161/CIRCULATIONAHA.121.055177.

33. Vemu PL, Yang E and Ebinger JE. Moving Toward a Consensus: Comparison of the 2023 ESH and 2017 ACC/AHA Hypertension Guidelines. JACC Adv 2024; 3: 101230. 2024/09/17. DOI: 10.1016/j.jacadv.2024.101230.

34. Stamatakis E, Owen KB, Shepherd L, et al. Is Cohort Representativeness Passe? Poststratified Associations of Lifestyle Risk Factors with Mortality in the UK Biobank. Epidemiology 2021; 32: 179–188. 2021/01/26. DOI: 10.1097/EDE.0000000000001316.

35. Ekelund U, Tarp J, Steene-Johannessen J, et al. Dose-response associations between accelerometry measured physical activity and sedentary time and all cause mortality: systematic review and harmonised meta-analysis. BMJ 2019; 366: l4570. 20190821. DOI: 10.1136/bmj.l4570.

36. Carlson SA, Fulton JE, Pratt M, et al. Inadequate physical activity and health care expenditures in the United States. Prog Cardiovasc Dis 2015; 57: 315–323. 20140809. DOI: 10.1016/j.pcad.2014.08.002.

37. van Alten S, Domingue BW, Faul J, et al. Reweighting UK Biobank corrects for pervasive selection bias due to volunteering. Int J Epidemiol 2024; 53 2024/05/08. DOI: 10.1093/ije/dyae054.

38. Koemel NA, Biswas RK, Ahmadi MN, et al. Minimum combined sleep, physical activity, and nutrition variations associated with lifeSPAN and healthSPAN improvements: a population cohort study. eClinicalMedicine 2026; 92. DOI: 10.1016/j.eclinm.2025.103741.

39. Kraus WE, Powell KE, Haskell WL, et al. Physical Activity, All-Cause and Cardiovascular Mortality, and Cardiovascular Disease. Med Sci Sports Exerc 2019; 51: 1270–1281. 2019/05/17. DOI: 10.1249/mss.0000000000001939.

40. Bakker EA, Lee D-c, Hopman MT, et al. Dose–response association between moderate to vigorous physical activity and incident morbidity and mortality for individuals with a different cardiovascular health status: A cohort study among 142,493 adults from the Netherlands. PLoS medicine 2021; 18: e1003845.

41. Lopez JPR, Gebel K, Chia D, et al. Associations of vigorous physical activity with all-cause, cardiovascular and cancer mortality among 64 913 adults. BMJ Open Sport & Exercise Medicine 2019; 5.

42. Pescatello LS, Franklin BA, Fagard R, et al. American College of Sports Medicine position stand. Exercise and hypertension. Med Sci Sports Exerc 2004; 36: 533–553. 2004/04/13. DOI: 10.1249/01.mss.0000115224.88514.3a.

43. Ekelund U, Tarp J, Steene-Johannessen J, et al. Dose-response associations between accelerometry measured physical activity and sedentary time and all cause mortality: systematic review and harmonised meta-analysis. bmj 2019; 366.

44. Arnett DK, Blumenthal RS, Albert MA, et al. 2019 ACC/AHA Guideline on the Primary Prevention of Cardiovascular Disease: A Report of the American College of Cardiology/American Heart Association Task Force on Clinical Practice Guidelines. Circulation 2019; 140: e596–e646. DOI: doi:10.1161/CIR.0000000000000678.

45. Ornish D, Scherwitz LW, Billings JH, et al. Intensive Lifestyle Changes for Reversal of Coronary Heart Disease. JAMA 1998; 280: 2001–2007. DOI: 10.1001/jama.280.23.2001.

46. Pescatello LS, Franklin BA, Fagard R, et al. Exercise and Hypertension. Medicine & Science in Sports & Exercise 2004; 36: 533-553. DOI: 10.1249/01.Mss.0000115224.88514.3a.

47. Cornelissen VA and Smart NA. Exercise training for blood pressure: a systematic review and meta-analysis. J Am Heart Assoc 2013; 2: e004473. 2013/03/26. DOI: 10.1161/jaha.112.004473.

48. Health UDo and Services H. 2018 Physical activity guidelines advisory committee scientific report. 2018: Part F. Chapter 6.

49. Powell KE, King AC, Buchner DM, et al. The Scientific Foundation for the Physical Activity Guidelines for Americans, 2nd Edition. Journal of Physical Activity and Health 2019; 16: 1–11. DOI: 10.1123/jpah.2018-0618.

50. Artinian NT, Fletcher GF, Mozaffarian D, et al. Interventions to Promote Physical Activity and Dietary Lifestyle Changes for Cardiovascular Risk Factor Reduction in Adults. Circulation 2010; 122: 406–441. DOI: doi:10.1161/CIR.0b013e3181e8edf1.

51. Bridger Staatz C, Gutin I, Tilstra A, et al. Midlife health in Britain and the United States: a comparison of two nationally representative cohorts. International Journal of Epidemiology 2024; 53. DOI: 10.1093/ije/dyae127.

52. Banks J, Marmot M, Oldfield Z, et al. Disease and Disadvantage in the United States and in England. JAMA 2006; 295: 2037–2045. DOI: 10.1001/jama.295.17.2037.

53. Mainous AG, 3rd, Diaz VA, Saxena S, et al. Diabetes management in the USA and England: comparative analysis of national surveys. J R Soc Med 2006; 99: 463–469. 2006/09/02. DOI: 10.1177/014107680609900918.

54. Desai M, Rachet B, Coleman MP, et al. Two countries divided by a common language: health systems in the UK and USA. J R Soc Med 2010; 103: 283–287. 2010/07/03. DOI: 10.1258/jrsm.2010.100126.

55. Papanicolas I, Mossialos E, Gundersen A, et al. Performance of UK National Health Service compared with other high income countries: observational study. BMJ 2019; 367: l6326. DOI: 10.1136/bmj.l6326.

56. Sallis JF, Cerin E, Conway TL, et al. Physical activity in relation to urban environments in 14 cities worldwide: a cross-sectional study. The Lancet 2016; 387: 2207–2217. DOI: 10.1016/S0140-6736(15)01284-2.

57. Prince SA, Adamo KB, Hamel ME, et al. A comparison of direct versus self-report measures for assessing physical activity in adults: a systematic review. Int J Behav Nutr Phys Act 2008; 5: 56. 20081106. DOI: 10.1186/1479-5868-5-56.

58. Dyrstad SM, Hansen BH, Holme IM, et al. Comparison of self-reported versus accelerometer-measured physical activity. Med Sci Sports Exerc 2014; 46: 99–106. 2013/06/25. DOI: 10.1249/MSS.0b013e3182a0595f.

59. Gropper H, John JM, Sudeck G, et al. The impact of life events and transitions on physical activity: A scoping review. PloS one 2020; 15: e0234794.

